# Epidemiological Modeling of Seasonal Influenza Through Periodic Mean-Reverting Stochastic Processes

**DOI:** 10.64898/2026.11.05.26367374

**Authors:** Fahad Mostafa, Linda J. S. Allen

**Affiliations:** School of Mathematical & Natural Sciences, Arizona State University, Phoenix, AZ 85051, USA; Department of Mathematics & Statistics, Texas Tech University, Lubbock, TX 79409, USA

**Keywords:** Data imputation, Seasonality, Stochastic models, Parameter estimation, Confidence region

## Abstract

Seasonal influenza follows annual cycles, but variation in epidemic magnitude and timing complicates preparedness. We applied periodic mean-reverting stochastic differential equations (SDEs) to seven seasons of weekly Texas influenza surveillance data. Missing observations were imputed using MissForest and compared with four alternative methods. Stochastic variability was modeled as *σpX*(*t*)^*p*^ *dW* (*t*), with *p* fixed at 1*/*2, 2*/*3, 5*/*6, and 1, representing square-root, intermediate sublinear, and multiplicative diffusion. Each specification was evaluated for direct influenza modeling and within a susceptible–exposed–infectious–recovered model incorporating waning immunity, where the SDE governed transmission. Parameters were estimated using least squares and approximate maximum likelihood for direct models and sample-proportion weighted least squares for stochastic SEIR models. Model-based parametric bootstrap assessed direct-model parameter uncertainty. For direct modeling, *p* = 1*/*2 achieved the lowest RMSE (712.8) and AIC (5383.0), with similar AIC for *p* = 2*/*3 (5383.2). For stochastic SEIR modeling, *p* = 1 achieved the lowest RMSE (0.02926), compared with 0.03506 for *p* = 1*/*2. The deterministic SEIR model yielded ℛ_0_ = 1.158. Transmission peaked approximately 10 weeks before infectious prevalence, while stochastic transmission generated variation in epidemic peak timing. Bootstrap analysis identified dependence between mean-reversion and diffusion parameters. Overall, the appropriate diffusion structure depended on the epidemiological quantity modeled, supporting flexible characterization of seasonal transmission variability while requiring prospective validation before operational forecasting.

## 1 Introduction

Seasonal influenza remains a major global public health challenge, causing substantial morbidity, mortality, and healthcare burden each year [2, 26]. Influenza epidemics exhibit strong annual periodicity, with predictable seasonal peaks driven by climatic conditions, host behavior, viral evolution, and population immunity, e.g., [47, 54, 73]. Despite these recurring patterns, epidemic timing, magnitude, and duration vary considerably because of stochastic environmental, demographic, and epidemiological factors. Accurate characterization of both seasonal structure and random variation is therefore essential for understanding disease transmission, improving outbreak forecasting, evaluating intervention strategies, and informing public health preparedness. Statistical and mechanistic models that jointly capture periodicity and stochasticity provide a robust framework for influenza surveillance and epidemiological inference.

Analyzing complex periodic data is essential for identifying and understanding recurring patterns in time series data, providing valuable insights into trends, seasonality, and anomalies [27]. Effective mathematical models for simulating systems that naturally exhibit randomness over time are stochastic differential equations (SDEs). SDEs have a long history in finance and economics, e.g., [13, 21, 42, 51, 65]. More recently, they been applied to ecology, epidemiology, and physiology [4, 30, 34, 35, 52, 58]. Mean-reverting SDE models that account for seasonal fluctuations are common in financial economics, energy markets, and commodity price modeling, e.g., [28, 29, 61, 53]. Also, mean-reverting SDEs with fluctuations in the mean reversion level have been fit directly to temperature and seasonal influenza data, e.g., [54, 78].

Seasonal disease forecasting relies on recognizing periodic patterns, most notably annual or subannual cycles driven by climate, host behavior, and pathogen biology, which shape predictable peaks such as winter influenza or rainy-season malaria [6, 31, 68]. However, it is even more critical to address stochasticity, since random fluctuations in weather, human behavior, pathogen evolution, and reporting noise can dramatically alter outbreak timing and size [62]. Ignoring this randomness leads to overconfident predictions, whereas stochastic models provide more realistic forecasts with uncertainty bounds, making them far more useful for guiding public health decisions.

A variety of statistical and mathematical approaches have been used to model seasonal influenza dynamics. Important in the statistical approaches are the inclusion of data to test reliability of modeling outcomes. Discrete time-series models such as a seasonal autoregressive integrated moving average model capture autocorrelation and seasonal variation in weekly influenza incidence [14, 38]. Inclusion of exogenous predictors, such as meteorological variables, Google Trends, and mobility data, improve forecasting accuracy [80]. Although computationally efficient for short-term prediction, these approaches remain primarily descriptive and lack epidemic stages [14]. Structural and state-space models extend classical time-series methods by distinguishing disease processes from noisy observations [25]. Dynamic linear and Bayesian state-space models have been widely applied to influenza surveillance, accommodating measurement error, missing data, and multiple surveillance streams [36, 37, 75], with Kalman and particle filtering enabling efficient real-time inference [66]. Likewise, Bayesian methods provide a framework for uncertainty quantification and data integration, supporting hierarchical surveillance models, transmission parameter estimation, and probabilistic epidemic forecasting [2, 15, 64, 76]. Because influenza incidence is observed as count data, generalized linear models (GLMs) are also widely used to estimate disease burden, characterize seasonality, and assess environmental and demographic risk factors [17, 36, 50]. However, these discrete-time methods do not model epidemic dynamics explicitly.

Many deterministic and stochastic mathematical models have captured some of the complex temporal and spatial dynamics of seasonal influenza in specific settings, e.g. [7, 10, 12, 16, 24, 26, 40, 47, 49, 57, 69, 73, 77, 79]. The majority of these epidemic models have an underlying Susceptible, Exposed, Infectious, and Recovered (SEIR) structure that include important characteristics of seasonal influenza spread and prevention (seasonal variability, waning immunity, multiple circulating viral types/subtypes, and vaccination). Some of the earliest mathematical investigations on seasonal infectious diseases applied SIR ordinary differential equations (ODEs) with a periodic transmission rate to study recurrent outbreaks of childhood diseases [9, 23]. A review of seasonal modeling studies by Altizer et al. [6] provides a comprehensive review of models for the spread of human and animal infectious diseases and presents some of the challenges in defining the seasonal drivers. To our knowledge, there has been no epidemic modeling applications using seasonal surveillance data to estimate parameters for periodic mean-reverting SDEs in epidemic models. However, a few studies have applied constant mean-reverting SDEs in parameters for disease transmission, death rate, or vector emergence in SEIR/SIR or epidemiological vector-host models to verify compatibility with routinely collected surveillance data [16, 49, 56, 63, 69].

Estimation of the mean-reverting parameters from discrete observations is important and can be facilitated by various numerical methods such as Euler-Maruyama and likelihood-based approaches [3, 39, 41, 72]. Other estimation methods include method of moments [3, 33] and filtering [55]. All of these methods are computationally intensive as they require a large number of numerically simulated sample paths. Among the most well-known mean-reverting diffusion process is the Ornstein–Uhlenbeck (OU) process, which models random fluctuations around a long-term equilibrium with a Gaussian distributed density [42].

Mean-reverting SDEs provide a useful stochastic framework for modeling epidemic dynamics as well as seasonal variability and long-term behavior, making them particularly well suited for studying influenza dynamics beyond short-term forecasting. Also, the time-continuous mean-reverting process may account for temporal correlation in social behavior of individuals [49]. In the stochastic SIR/SEIR literature, periodic mean-reverting processes have been largely unexplored. Only in a simplified setting, without the epidemic structure, were parameters for mean-reversion and volatility fit directly to influenza data [54]. Therefore, our primary goals are to fill this gap, to provide a foundational study in epidemiological modeling by extending this research to several new seasonal mean-reverting processes, formulating a new SEIR compartmental model with seasonal mean-reverting transmission, and fitting the mean-reversion and volatility parameters of these models to seasonal influenza data. In the next section, we define and discuss the properties of the periodic mean-reverting SDEs.

## 2 Periodic Mean-Reverting SDEs

We consider the following Itô periodic mean-reverting SDEs:

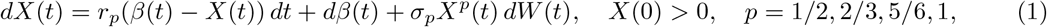

where *t* ∈ [0,∞), *r*_*p*_, *σ*_*p*_ *>* 0, and *β*(*t*) are positive, periodic functions. Parameter *r*_*p*_ is the mean-reversion rate and *σ*_*p*_ is the coefficient for the diffusion term *σ*_*p*_*X*^*p*^(*t*). The expectation of the SDEs in Equation (1) is given by

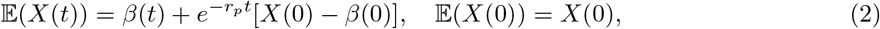

and therefore, the asymptotic mean equals *β*(*t*) [54]. We assume the periodic function *β*(*t*) is also continuous with a continuous derivative. If *β*(*t*) is a constant function 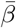, then the SDEs in Equation (1) are referred to as the Chan-Karolyi-Longstaff-Sanders family of models [42]. The two cases *p* = 1*/*2 and *p* = 1 are often referred to as the Cox-Ingersoll-Ross (CIR) process and geometric Brownian motion (GBM), respectively, e.g. [21, 42, 54]. These two cases have been studied in some detail in reference [54]. However, the combination of periodic mean *β*(*t*) and state-dependent diffusion *σ*_*p*_*X*^*p*^(*t*) and the roles played by parameters *r*_*p*_ and *σ*_*p*_ for *p* ∈ [1*/*2, 1] with respect to data have not been studied. The diffusion term *σ*_*p*_*X*^*p*^(*t*) *dW* (*t*), *p* = 1*/*2, 2*/*3, 5*/*6, 1, describes how stochastic variability changes with the underlying level of influenza activity *X*(*t*). Conditional on *X*(*t*) = *x*, the infinitesimal variance is

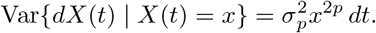

Thus, *p* determines the degree of state-dependent heteroscedasticity. For *p* = 1*/*2, variability is consistent with approximately event-level or demographic fluctuations, with variance proportional to influenza activity. The intermediate values *p* = 2*/*3 and *p* = 5*/*6 allow progressively stronger variability during periods of elevated influenza activity, potentially reflecting heterogeneous contacts, localized outbreaks, behavioral changes, or environmental perturbations. For *p* = 1, the diffusion is fully multiplicative, so random fluctuations are proportional to the prevailing influenza level. The relative diffusion intensity,

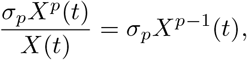

decreases with influenza activity when *p <* 1 but remains constant when *p* = 1. Hence, increasing *p* represents a transition from approximately event-driven variability toward increasingly multiplicative epidemic uncertainty. In this study, *p* is fixed *a priori* at each candidate value rather than estimated continuously, and the remaining parameters are estimated separately for each specification. In the results section, the influence of *p* is discussed further.

We do not consider the OU process with *p* = 0 which has been widely used in stochastic modeling because of its tractable Gaussian distribution. A key limitation of the OU process is that its stationary distribution is normal with unbounded support, which means the process does not remain strictly positive and can cross into negative values even when centered around a positive mean [22]. This feature makes the OU process unsuitable in settings where the underlying quantity is naturally nonnegative.

Nonnegative solutions are an important property in biological systems. If 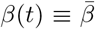 is constant with *p* ∈ (1*/*2, 1], then *X*(*t*) *>* 0 for *t >* 0 and if *p* = 1*/*2, then *X*(*t*) ≥ 0 for *t >* 0 [42]. The SDEs in Equation (1) with positive and periodic *β*(*t*) and the state-dependent diffusion terms have nonnegative solutions provided *r*_*p*_*β*(*t*) + *β*^′^(*t*) ≥ 0 [5]. To ensure the numerical methods generate nonnegative sample paths, we set *X*(*t*) = 0, if *X*(*t*) ≤ 0 for *t >* 0.

The parameters of the four SDEs in Equation (1) are fit to seven years of weekly seasonal influenza data as described in the next section.

The range of power values *p* = 1*/*2, 2*/*3, 5*/*6, 1 allow us to explore how subtle changes in diffusion behavior impact model fitting and parameter uncertainty, particularly under seasonal fluctuations represented by the periodic function *β*(*t*). Our bootstrap-based confidence regions reflect how sensitive parameter estimation is to this nonlinear structure, and help distinguish between models with similar drift but differing stochastic dynamics.

In the next section, the seasonal influenza data and missing data imputation methods are discussed. In Section 4, parameters of the periodic mean-reverting SDEs in Equation (1) are fit to the imputed influenza data for each case *p* ∈ *{*1*/*2, 2*/*3, 5*/*6, 1} and parameter uncertainty is studied with a bootstrap analysis and the results interpreted. In Section 5, the new SEIR SDE model with a periodic mean-reverting transmission rate is discussed. The model is fit to the imputed influenza data for the four cases *p*∈ *{*1*/*2, 2*/*3, 5*/*6, 1} and the epidemiological implications reported. The final section is a summary and discussion of the results, the model limitations, and the areas for future research.

## 3 Seasonal Influenza Data

### 3.1 Machine Learning for Missing Data Imputation

Weekly influenza case data (number of specimens tested) were collected from Texas Influenza Surveillance Activity Reports from the 2012–2013 to 2018–2019 seasons beginning at week 40 [71]. According to the Texas Department of State Health Services (DSHS), the state is divided into 11 Health Service Regions. The Infectious Disease Control Unit of the DSHS, in collaboration with the DSHS Medical Virology Laboratory, monitors influenza activity across the state [71]. Participating healthcare providers collect and submit specimens weekly during the influenza season from patients exhibiting influenza-like illness symptoms. Our dataset comprises seven full influenza years, beginning from 2012-2013 and ending with the 2019–2020 season to avoid potential influences from COVID-19 in 2020. Each year is assumed to follow a 52-week cycle, with some overlap between consecutive years.

Several missing data imputation techniques are evaluated, namely, Multiple Imputation by Chained Equations (mice), Piecewise Cubic Hermite Interpolating Polynomial (pchip), Cubic Spline Interpolation (spline), Modified Akima Interpolation (makima), and MissForest [1, 46, 48, 54]. We assess the performance using root mean square error (RMSE) against the mean imputed data points of the original data. The missing value imputation is formulated in a periodic pattern. In this context, let *Z* denote the weekly time-to-event case matrix over seven years. For each year, *s* ∈ {1, …, 7}, define the set of observed weeks as 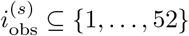 and the set of missing weeks as 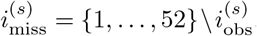. The corresponding observed responses are 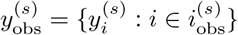 and the missing responses are 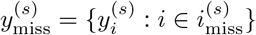. Let the time indices for observed and missing entries be denoted as

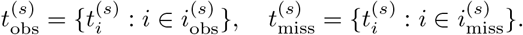

The mice method iteratively models each variable with missing values as a conditional distribution given all other variables. For each *s*, the regression model is denoted as

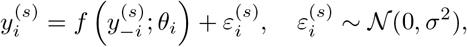

where *f* is typically least square regression, and 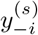 denotes all other observed variables. The model is updated iteratively, imputing missing entries from the posterior predictive distribution. However, the pchip method constructs a piecewise cubic polynomial *g*^(*s*)^(*t*) such that

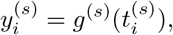

and the first derivative is continuous and preserves monotonicity. The values 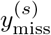 are then found by evaluating *g*^(*s*)^ 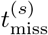.

The cubic spline interpolation fits a smooth function *h*^(*s*)^(*t*) such that

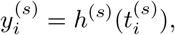

with continuity up to the second derivative. The imputed values are 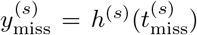. The makima method also constructs a piecewise function *a*^(*s*)^(*t*) using local slopes to reduce oscillations. It is more robust near sharp gradients, using a weighted average of neighboring slopes,

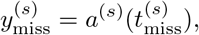

with *a*^(*s*)^(*t*) derived from piecewise rational linear expressions based on 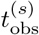 and 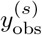.

The MissForest method is a nonparametric approach that models missing values using an ensemble of decision trees. In this case, a random forest regressor 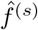 is trained on the observed time-response pairs, i.e., 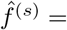 MissForest 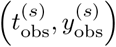. This model is then used to impute the missing values as

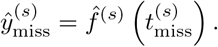

The above imputation is repeated iteratively (e.g., 100 iterations) for each year *s* = 1, …, 7. The final imputed case matrix *Z*, is then formed for each of the imputation methods. For the MissForest method,

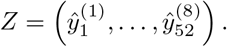

To compare the results of the five imputation methods, the RMSE is used to evaluate the average deviation between actual and predicted values, 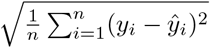, where *y*_*i*_ represents mean imputed data points of the original data, *ŷ*_*i*_ denotes the imputed values, and *n* is the total number of data points. Figure 1 presents the comparison of the RMSE across the five imputation methods, where each method is evaluated over 1000 bootstrap replicates using perturbed versions of the imputed variables to account for variability and to reflect realistic uncertainty in reconstruction. The results demonstrate that while interpolation-based methods (pchip, spline, and makima) and the multiple imputation approach mice yield RMSE values in a similar range (approximately 9.0 to 10.5), the MissForest algorithm achieves significantly lower RMSE values, with a median close to 1.0. This stark contrast indicates that MissForest, which leverages random forests to model complex nonlinear interactions in the case of periodic data patterns, provides a markedly more accurate imputation of the missing data. Moreover, these findings are consistent with the literature emphasizing MissForest’s ability to model nonlinear, complex data patterns more effectively than traditional interpolation and regression-based imputations [70]. Furthermore, the imputation comparison indicates that MissForest provides substantially lower RMSE than other models. The RMSE was evaluated against artificially removed observations with known values, ensuring direct assessment of reconstruction accuracy [67]. Nevertheless, machine-learning-based imputation may smooth local fluctuations and potentially influence subsequent stochastic modeling. Therefore, alternative imputation methods were considered as robustness checks. Comparing the resulting stochastic parameter estimates and epidemic characteristics across imputation approaches helps determine whether the main conclusions are robust to the choice of missing-data reconstruction method.

**Figure 1:**
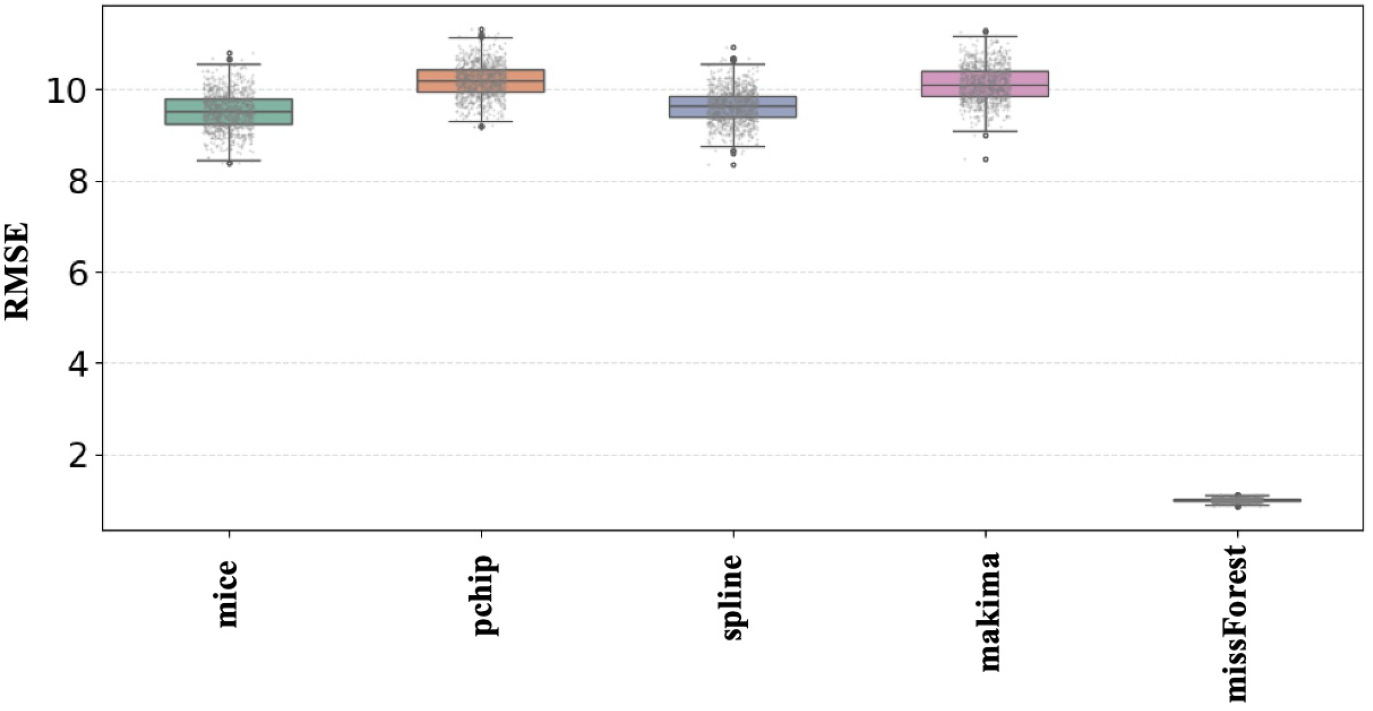
RMSE comparison across multiple imputation methods for the influenza dataset. Lower RMSE indicates greater accuracy in reconstructing the artificially masked observations. The MissForest algorithm yielded the lowest RMSE among the evaluated imputation methods.

The imputation framework applied in this investigation is implemented using the MissForest algorithm as the RMSE indicates it has the greatest accuracy. Figure 9 in Appendix A is an illustration of the imputation of missing weekly influenza cases for the MissForest algorithm.

## 4 Mean-Reverting SDEs Applied to Influenza

### 4.1 Point Estimation of Parameters

The first objective is to find point estimations of the parameters associated with the mean-reverting SDEs in Equation (1) based on the time-series data 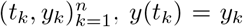. We apply techniques from our previous research in [54] to fit two additional SDEs in Equation (1), *p* = 2*/*3, 5*/*6 and to calculate the RMSE. These methods are summarized below; more details can be found in [54].

First, the data are fit to a smooth periodic function *β*(*t*) with period equal to one year, *β*(*t*) = *β*(*t* + 1) (E(*X*(*t*)) = *β*(*t*)). The function is a sum of scaled normal-type distributions with three parameters, *a*_1_, *a*_2_, *a*_3_:

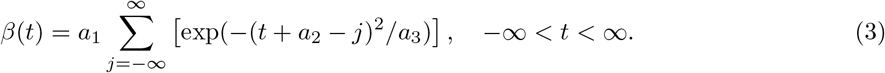

We tested a sinusoidal function, but the fit resulted in negative values; therefore, we chose this new function (3) to ensure positivity and parsimony. The infinite series can be approximated well by a finite series because the data consists of seven years, *t* ∈ [0, 7]. The parameters *a* = (*a*_1_, *a*_2_, *a*_3_) are estimated by minimizing the sum of squared errors,

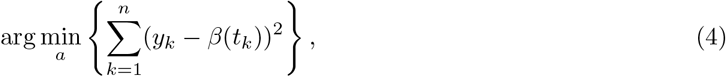

where *n* is the number of data points. We also compute the RMSE. Second, the data are used to estimate the values of *r*_*p*_ and *σ*_*p*_ via a maximum likelihood method. The normal density and the Euler-Maruyama discretization are applied as in [3, 41]. Let *t* = *t*_*k*_ and *X*(*t*_*k*_) = *x*_*k*_. The periodic mean-reverting SDEs in Equation (1) are written in a discrete form:

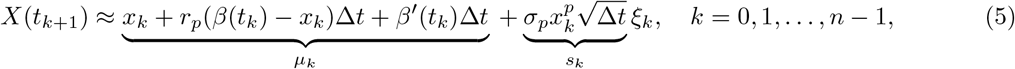

where *ξ*_*k*_ ~ *N* (0, 1). This leads to a transition probability with a normal density:

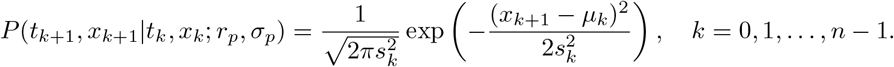

The parameters *r*_*p*_ and *σ*_*p*_ are found by minimizing the negative of the log-likelihood function.

To calculate the RMSE for the fit of the SDEs, Equation (5) is applied. Let 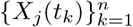 denote the predicted values or sample path values at time *t*_*k*_ from the SDE model with the estimated parameters *r*_*p*_ and *σ*_*p*_ and those for *β*(*t*) with Δ*t* = *t*_*k*+1_ − *t*_*k*_ (duration one week for the influenza data). Therefore,

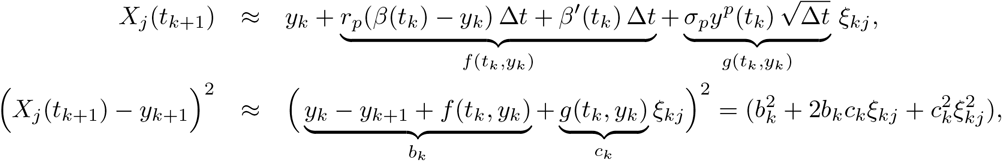

where *ξ*_*jk*_ ~ *N* (0, 1). Summing the squared errors, averaging over a large number (*N*) of sample paths and applying the approximations, 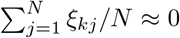 and 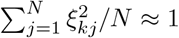, lead to a simple expression of the RMSE for the SDEs in Equation (1):

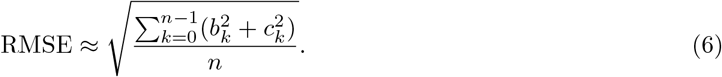

The point estimations for the three parameters in *β*(*t*), *a*_1_, *a*_2_, and *a*_3_, the two parameters in the SDEs, *r*_*p*_, and *σ*_*p*_, and their respective RMSEs are summarized in Table 1. The Akaike Information Criterion (AIC) is also reported for the joint estimation of the mean-reversion parameter *r*_*p*_ and the diffusion parameter *σ*_*p*_. The AIC provides a likelihood-based measure of model fit while penalizing model complexity, thereby facilitating comparison among alternative diffusion specifications indexed by *p*. Lower AIC values indicate greater empirical support for a particular specification, reflecting a more favorable balance between goodness of fit and model complexity. The condition *r*_*p*_*β*(*t*) + *β*^′^(*t*) ≥0 that guarantees nonnegative solutions of the SDEs are satisfied for these particular parameter values.

**Table 1:**
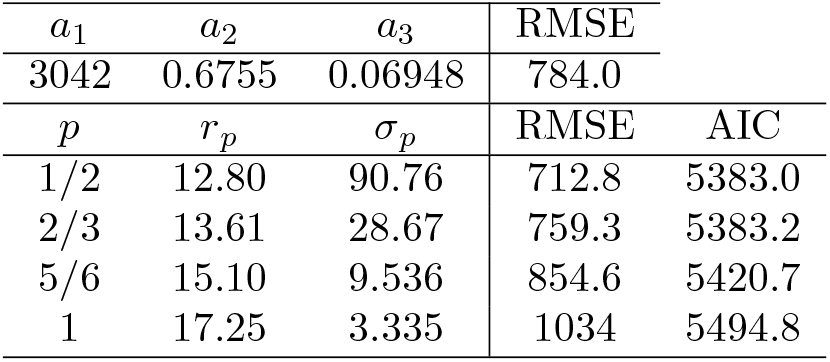
Point estimates of the five model parameters in Equations (1) and (3) and values for the RMSE and AIC.

The top row of the Table 1 reports point estimates for parameters *a*_1_, *a*_2_, and *a*_3_, yielding an RMSE of 784.0. The lower panel shows estimates of *r*_*p*_ and *σ*_*p*_ for fixed *p*∈ *{*1*/*2, 2*/*3, 5*/*6, 1}, where increasing *p* leads to larger *r*_*p*_ and smaller *σ*_*p*_. The cases 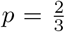 and 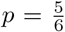 illustrate a transition between squareroot-type diffusion and fully linear diffusion in terms of model behavior and fit quality. For 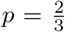, the model achieves a relatively low RMSE of 759.3 (AIC=5383.2), indicating good overall fit. The volatility estimate *σ*_2*/*3_ = 28.67 is significantly lower than the square-root case (*σ*_1*/*2_ = 90.76) but still substantially higher than in the more linear regimes. The corresponding mean-reversion rate *r*_2*/*3_ = 13.61 suggests a moderately strong pull of the process toward the seasonal mean *β*(*t*). This setting reflects a model where diffusion remains sensitive to the current state *X*(*t*), but without the extreme volatility amplification seen in the square-root case. For 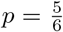, the estimated volatility drops further to *σ*_5*/*6_ = 9.536, with a higher mean-reversion rate of *r*_5*/*6_ = 15.10. While the RMSE increases to 854.6, the diffusion term becomes more state-dependent, but still sublinear, preserving the qualitative properties of the model without becoming overly rigid or explosive.

Plots of the influenza case data, the fit of the period mean *β*(*t*), and a sample path of the SDEs for the two cases *p* = 1*/*2, 1 over seven years are shown in Figure 2. In the left panel, the close agreement between the data mean and periodic mean *β*(*t*) can be seen as well as some variability near the peak of the influenza data (data mean+SD). The fit of the mean *β*(*t*) indicates a dominant contribution from the amplitude *a*_1_ due to the large data spikes in annual cases during the winter season (Table 1). A large decrease in the volatility parameter *σ*_*p*_ and a slight increase of the mean-reversion rate *r*_*p*_ is seen as the value of *p* increases. Also, the maximum peaks of the sample paths increase when *p* = 1 as compared to *p* = 1*/*2 (Figure 2). The RMSE has the smallest value when *p* = 1*/*2 and the largest when *p* = 1, highlighting a trade-off between *p* and the fit, i.e., the fit is reduced with the larger value of *p* (Table 1). This suggests that stochastic effects through the diffusion term *σ*_*p*_*X*^*p*^(*t*) are critical for accurately modeling system dynamics.

**Figure 2:**
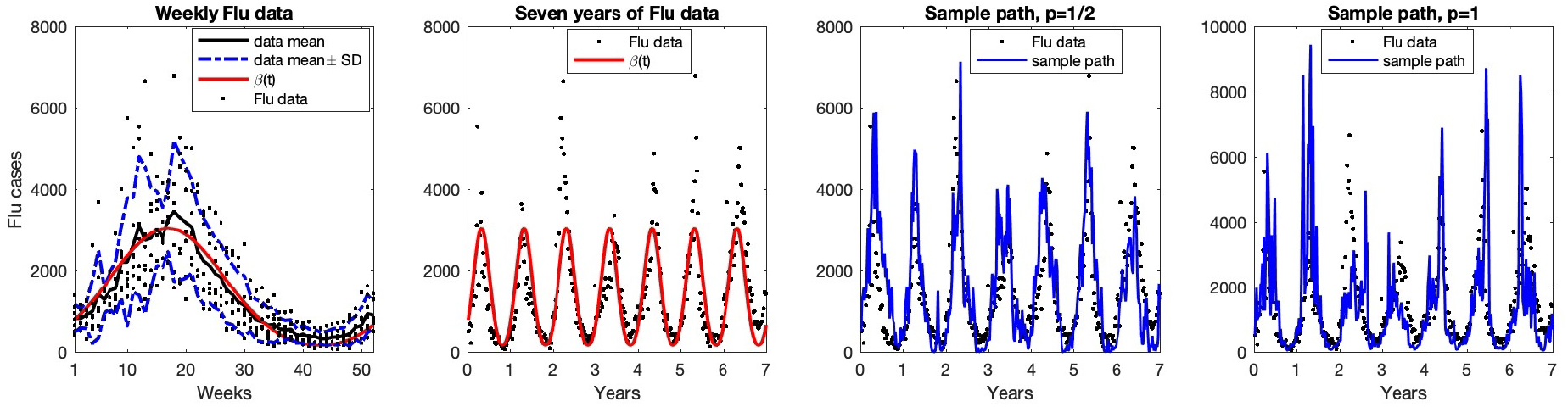
In the left panel, seven years of weekly seasonal influenza data (52 weeks/year) with the data mean (black curve), data mean ± SD (standard deviation) (dashed blue curves) and the periodic mean *β*(*t*) (red curve) are graphed over 52 weeks. In the second panel, the influenza data and *β*(*t*) (red curve) are graphed over seven years. In the last two panels, the curve *β*(*t*) (red curve) and a sample path for each of the SDEs in Equation (1) (blue curves) for *p* = 1*/*2, 1, respectively, are graphed over seven years.

### 4.2 Confidence Ellipses for Model Parameters

Assuming that the SDEs in Equation (1) are the data-generating models for the original data, we apply the parameter estimates in Table 1 to test these models. One hundred sample paths for seven years are generated from each of the four SDEs in Equation (1) (*p* = 1*/*2, 2*/*3, 5*/*6, 1) to obtain 100 estimates for the five model parameters.

Parameter uncertainty was quantified using a model-based parametric bootstrap designed to preserve the temporal dependence induced by the fitted SDE. Let 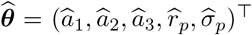 denote the parameter vector estimated from the observed time series. Conditional on ***θ***, we generated *B* = 100 independent bootstrap trajectories for *T* = 7 years

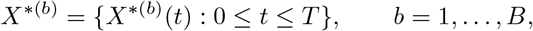

from the fitted SDE model. Thus, the bootstrap is based on simulated stochastic paths rather than on resampling individual residuals or observations. For each bootstrap replication *b*, the initial condition was randomly selected from the corresponding annual starting values of the observed series,

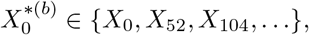

thereby allowing uncertainty associated with the initial state to contribute to the bootstrap distribution. Conditional on this initial value, a complete seven-year trajectory was generated using Milstein’s numerical scheme [3] with integration step Δ*t* = 1*/*520. The simulated trajectory was subsequently sampled at weekly intervals, *t*_*j*_ = *j/*52, *j* = 0, …, 364, so that each bootstrap dataset had the same sampling frequency and duration as the observed series. Importantly, observations within a bootstrap series were not independently resampled. Instead, each trajectory was generated recursively from the fitted stochastic process. Consequently, the serial dependence, periodic mean structure, mean-reversion mechanism, and diffusion variability specified by the fitted SDE were propagated through the complete bootstrap trajectory. This model-based construction is preferable to an ordinary independent bootstrap for temporally dependent observations because it retains the dependence structure implied by the stochastic model. For each bootstrap dataset *X*^*(*b*)^, the complete parameter-estimation procedure used for the original data was repeated, producing

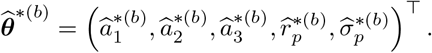

The empirical bootstrap mean was calculated as

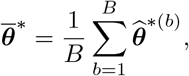

and the corresponding bootstrap covariance matrix was estimated by

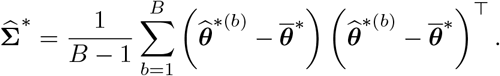

To quantify joint uncertainty between pairs of parameters, consider **Θ**_*jk*_ = (*θ*_*j*_, *θ*_*k*_)^⊤^, with bootstrap Mean 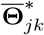 and the corresponding 2 × 2 submatrix 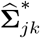 of 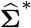. Under the bivariate-normal approximation to the bootstrap sampling distribution, the squared Mahalanobis distance is

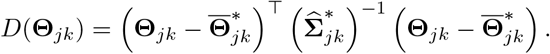

The 100(1 − *α*)% joint confidence region is therefore

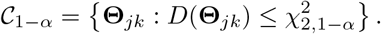

The region *D*(Θ) ≤*D*_{1−*α*,2}_ = 5.991 defines the 95% confidence ellipse at a significance level *α* = 0.05, [60].

Moreover, the construction of the bootstrap confidence ellipses assumes that the joint bootstrap distribution of each parameter pair is approximately bivariate normal with a finite and nonsingular covariance matrix. This assumption was assessed using parameter-pair scatterplots and Q–Q plots of the bootstrap Mahalanobis distances against the 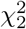 distribution. Because complete trajectories were generated from the fitted SDE rather than independently resampling individual observations, the model-implied temporal dependence, stochastic variability, and periodic structure were preserved within each bootstrap replicate.

Confidence ellipses are constructed for the parameter pairs (*r*_*p*_, *σ*_*p*_), (*a*_1_, *a*_3_), (*a*_1_, *a*_2_), and (*a*_3_, *a*_2_) in Figure 3. The four rows (A), (B), (C), and (D) correspond to the four cases *p* = 1*/*2, 2/3, 5/6, and 1, respectively. Their orientation and eccentricity provide a graphical representation of the bootstrap covariance between parameter estimates: nearly circular regions indicate weak dependence, whereas elongated ellipses indicate stronger joint estimation uncertainty or correlation.

**Figure 3:**
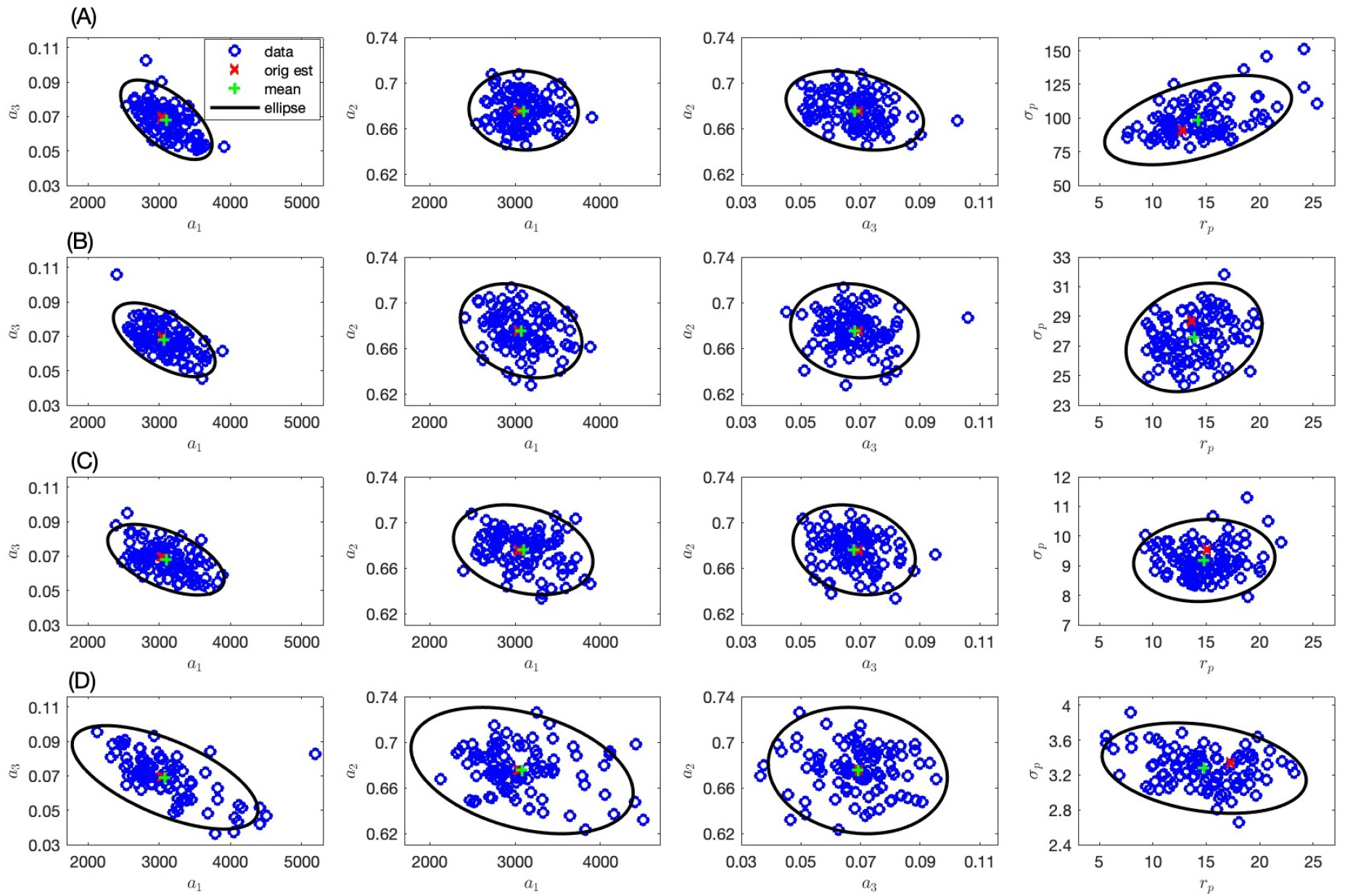
Plotted are one hundred bootstrap parameter estimates and 95% confidence ellipses for parameter pairs based on the original estimates in Table 1. For panels in row (A) *p* = 1*/*2, (B) *p* = 2*/*3, (C) *p* = 5*/*6, and (D) *p* = 1. The red cross and green star within each ellipse represent the original estimate of the parameter pair and the average of the bootstrap estimates, respectively.

### 4.3 Results

The uncertainty in parameter estimation through bootstrap resampling is illustrated in Figure 3. For each value of *p*, 100 bootstrap samples were used to re-estimate the parameters. The scatter of the blue circles shows the sampling variability. The red cross denotes the original parameter estimate (based on the full data), while the green star represents the average of the bootstrap estimates. The black ellipse encloses approximately 95% of the bootstrap estimates, giving a visual representation of the confidence region. Moreover, in Figure 3, each row corresponds to a different value of the parameter *p*. As *p* increases from 1*/*2 to 1, the shape and size of the confidence ellipses change, indicating the effect of *p* on the stability and precision of the parameter estimates. Notably, in panel (D) where *p* = 1, the confidence regions appear larger and more elongated for some parameter pairs, suggesting increased uncertainty or correlation among parameters.

The sample statistics from the bootstrap parameter estimates are summarized in Table 2. Parameters *a*_1_, *a*_2_, and *a*_3_ control the amplitude, time shift, and width of the peaks of the mean *β*(*t*), respectively. Parameters *a*_1_ and *a*_2_ have the strongest negative correlation, large *a*_1_ (large amplitude) is associated with small *a*_2_ (small time shift). Strong negative correlations between the shape parameters *a*_1_, *a*_2_, and *a*_3_ persist across the values, suggesting a robust structural relationship in the model’s nonlinear component regardless of the noise scaling.

**Table 2:**
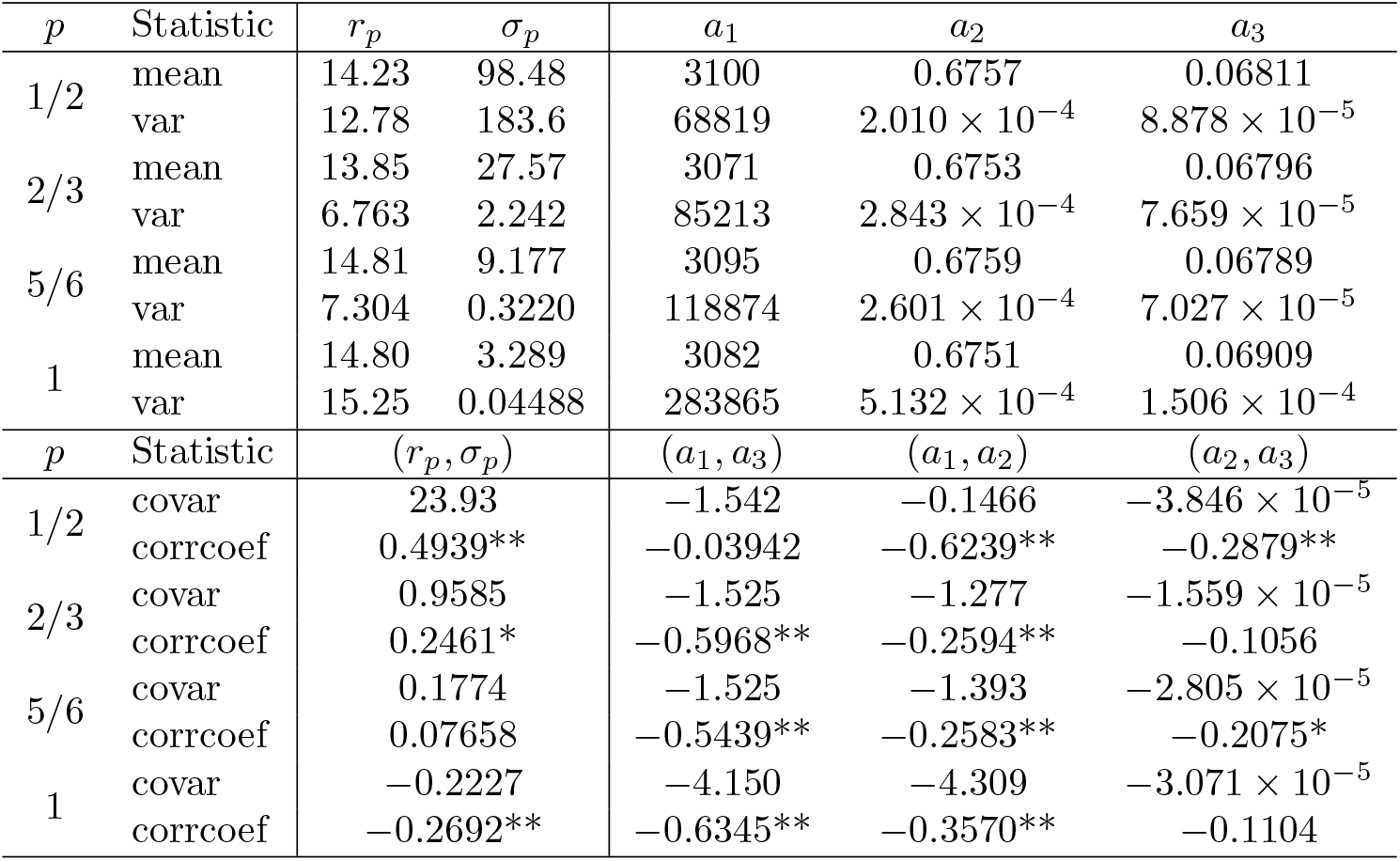
Summary statistics from the bootstrap data include the mean and variance (var) for all parameters and the covariance (covar) and correlation coefficient (corrcoef) for parameter pairs. Significant negative or positive correlation for the pairs indicated by * for 0.01 *< α <* 0.05 and ** for *α <* 0.01.

| *p* | Statistic | *r<sub>p</sub>* | *σ<sub>p</sub>* | *a*<sub>1</sub> | *a*<sub>2</sub> | *a*<sub>3</sub> |
|---|---|---|---|---|---|---|
| 1/2 | mean | 14.23 | 98.48 | 3100 | 0.6757 | 0.06811 |
|  | var | 12.78 | 183.6 | 68819 | 2.010 × 10<sup>−4</sup> | 8.878 × 10<sup>−5</sup> |
| 2/3 | mean | 13.85 | 27.57 | 3071 | 0.6753 | 0.06796 |
|  | var | 6.763 | 2.242 | 85213 | 2.843 × 10<sup>−4</sup> | 7.659 × 10<sup>−5</sup> |
| 5/6 | mean | 14.81 | 9.177 | 3095 | 0.6759 | 0.06789 |
|  | var | 7.304 | 0.3220 | 118874 | 2.601 × 10<sup>−4</sup> | 7.027 × 10<sup>−5</sup> |
| 1 | mean | 14.80 | 3.289 | 3082 | 0.6751 | 0.06909 |
|  | var | 15.25 | 0.04488 | 283865 | 5.132 × 10<sup>−4</sup> | 1.506 × 10<sup>−4</sup> |
| *p* | Statistic | (*r<sub>p</sub>*, *σ<sub>p</sub>*) |  | (*a*<sub>1</sub>, *a*<sub>3</sub>) | (*a*<sub>1</sub>, *a*<sub>2</sub>) | (*a*<sub>2</sub>, *a*<sub>3</sub>) |
| 1/2 | covar | 23.93 |  | −1.542 | −0.1466 | −3.846 × 10<sup>−5</sup> |
|  | corrcoef | 0.4939** |  | −0.03942 | −0.6239** | −0.2879** |
| 2/3 | covar | 0.9585 |  | −1.525 | −1.277 | −1.559 × 10<sup>−5</sup> |
|  | corrcoef | 0.2461* |  | −0.5968** | −0.2594** | −0.1056 |
| 5/6 | covar | 0.1774 |  | −1.525 | −1.393 | −2.805 × 10<sup>−5</sup> |
|  | corrcoef | 0.07658 |  | −0.5439** | −0.2583** | −0.2075* |
| 1 | covar | −0.2227 |  | −4.150 | −4.309 | −3.071 × 10<sup>−5</sup> |
|  | corrcoef | −0.2692** |  | −0.6345** | −0.3570** | −0.1104 |

The mean reversion parameter *r*_*p*_ accounts for return to the mean *β*(*t*). The estimates for the mean reversion rate *r*_*p*_ for all four SDE models (1) are comparable, as *r*_*p*_ values are measured in the same units, 1*/year*. The original parameter estimates for *r*_*p*_ in Table 1 increase from 12.8 to 17.25 as *p* increases but the estimates of *r*_*p*_ from the mean of the bootstrap parameter data in Table 2 remain fairly stable ranging from 13.85 to 14.81 with variance ranging from 6.763 to 15.25. This is an indication of a consistent mean-reversion force in the SDE models.

The volatility parameter *σ*_*p*_ affects the variability about the mean. The estimates of *σ*_*p*_ from the original data in Table 1 and from the mean of the bootstrap parameter estimates in Table 2 decrease significantly as *p* increases, e.g., dropping from 98.48 at *p* = 1*/*2 to 3.289 at *p* = 1 in Table 2. However, the estimates of *σ*_*p*_ are not strictly comparable because the units of *σ*_*p*_ depend on *p* and influenza cases as given by

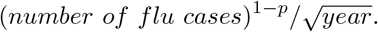

If the *σ*_*p*_ parameter is scaled using information from the original influenza data in Figure 2 so that *σ*_*p*_ have the same units of 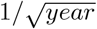 for all values of *p*, then the scaled *σ*_*p*_ values are stable. In particular, when the original parameter values *σ*_*p*_ in Table 1 are scaled by the median 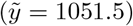 or the mean 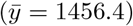 of the original influenza data, e.g. 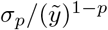, then the scaled *σ*_*p*_ remain reasonably stable, increasing from 2.80 to 3.335 for the median and from 2.38 to 3.335 for the mean as *p* increases from 1/2 to 1. Another method of comparison for the *r*_*p*_ and *σ*_*p*_ values as *p* increases is through the graphs of the reversion rates expressed in terms of the exponential density function 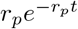 and the graphs of the volatility parameters expressed in terms of the diffusion coefficient *σ*_*p*_*X*^*p*^ (Figure 4). The exponential density functions show close agreement. The graphs of *σ*_*p*_*X*^*p*^ for all *p* = 1*/*2, 2*/*3, 3*/*4, 1 are close when *X* ∈ [0, 1500] for the original parameters and at *p* = 1*/*2 and *p* = 2*/*3 when *X* ∈ [0, 3000] for the mean bootstrap parameters. For larger values of *X*, there is greater disagreement between the graphs, especially for the linear case, *p* = 1.

**Figure 4:**
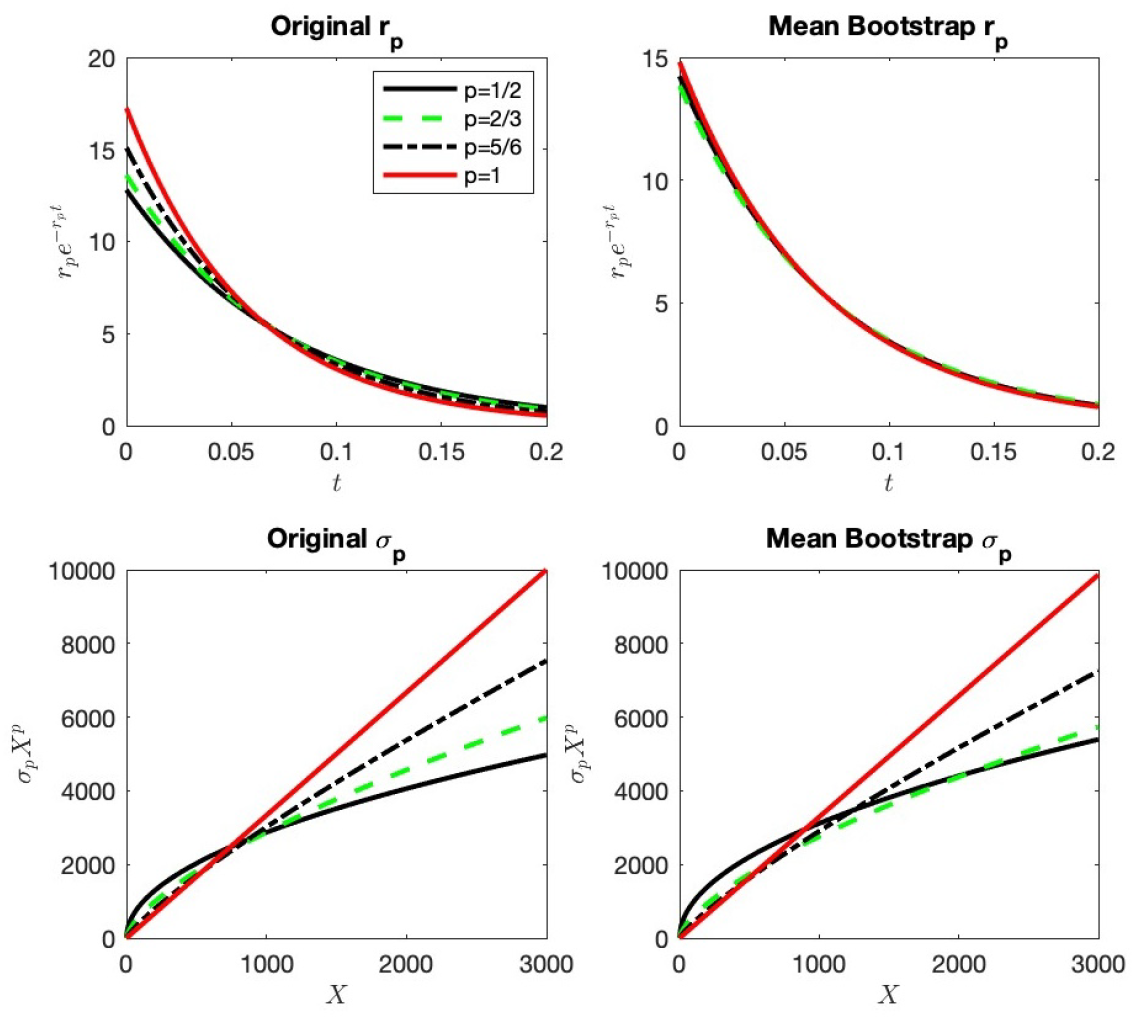
Graphed are the exponential density 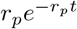 as a function of *t* (time in years) and the diffusion coefficient *σ*_*p*_*X*^*p*^ as a function of *X* (number of flu cases). Values of *r*_*p*_ and *σ*_*p*_ are given in Tables 1 and 2 for *p* = 1*/*2, 2*/*3, 5*/*6, 1.

The statistics in Table 2 provide information about correlation between the mean reversion rate *r*_*p*_ and the volatility parameter *σ*_*p*_. A significant positive correlation between *r*_*p*_ and *σ*_*p*_ (0.4939) is observed for *p* = 1*/*2, but this correlation weakens as *p* increases to 2/3 and 5/6 and becomes negative at *p* = 1 (−0.2692). The change in the correlation can be explained by considering the full nonlinear models, the drift term *r*_*p*_(*β*(*t*) −*X*(*t*))+*β*^′^(*t*), and the diffusion term *σ*_*p*_*X*^*p*^(*t*). These expressions have opposing effects in the SDE models; the drift *r*_*p*_(*β*(*t*) −*X*(*t*)) + *β*^′^(*t*) brings the dynamics toward the mean, whereas the diffusion *σ*_*p*_*X*^*p*^(*t*) allows the dynamics to grow and reach the high peaks in the seasonal data. The power *p* in the diffusion term has a large effect on the magnitude obtained by the sample paths with small values of *p* taking longer to reach the data peaks than large values of *p*. The volatility parameter alleviates this discrepancy. To compensate for small growth in the diffusion term with *p* = 1*/*2, small increases in *r*_*p*_ require large increases in the value of *σ*_*p*_ (positive correlation), whereas for large growth in the diffusion term with *p* = 1, small increases in *r*_*p*_ require large decreases in the value of *σ*_*p*_ (negative correlation).

## 5 Epidemic Model with Mean-Reverting Transmission

### 5.1 Stochastic Model in Epidemiology

The seasonal influenza data is used to fit a structured SEIR SDE model with a periodic mean-reverting transmission rate and waning immunity. The model does not include explicit spatial heterogeneity, multiple strains, vaccination, or environmental and social factors. But some of these factors are implicitly assumed in the model. Seasonal variability in an epidemic can be driven by environmental and social factors through a seasonal mean-reverting transmission rate, and a successful vaccination program can reduce the susceptible population size. The goals are to illustrate methods to fit the mean-reverting parameters in a simple but well-known SEIR model to the influenza data. The transmission rate, denoted as *Y* (*t*), is assumed to satisfy one of the four SDEs in Equation (1), *p* = 1*/*2, 2*/*3, 5*/*6, 1, making the full model a system of SDEs. The differential equations for the four SEIR stages and the transmission rate *Y* (*t*) yield the following system of SEIR SDEs:

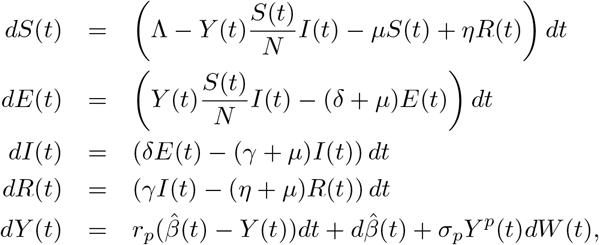

where parameter Λ is the birth/immigration rate, *µ* is the per capita natural death rate, *η* is the per capita rate of waning immunity, *δ* is the per capita transition rate from the exposed to the infectious state, and *γ* is the per capita recovery rate.

The periodic function 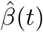 plays a different role in the SEIR model than the function *β*(*t*) in the SDE Equation (1). In the SEIR model, 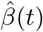 represents the periodic mean of the transmission rate with units per year, whereas *β*(*t*) in the SDE (1) is the periodic mean of the influenza cases. Therefore, the mean transmission rate 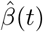 cannot be fit directly to the influenza data. Instead we fit the dynamic variable *I*(*t*) in the differential equations to the influenza data.

First, we make some simplifying and epidemiologically realistic assumptions about the SEIR demo-graphic and epidemiological parameters: population size *N* = 300, 000, death rate *µ* = 1*/*76*/yr*, transition rate from the exposed to infectious stage *δ* = 52*/yr*, recovery rate *γ* = 104*/yr*, waning immunity rate *η* = 4*/yr*, and birth/immigration rate Λ = *µN* = (300, 000*/*76)*people/yr*. Several parameters values were first tested within a reasonable range to obtain the best fit (Section 5.2). With these parameter values, the population size is constant at the disease-free state and the average life expectancy is 76 years. In the SEIR model, the average duration of immunity is assumed to be 3 months, the average duration for the exposed (or latent) period 7 days, and the average duration of the infectious period 3.5 days. Estimates for seasonal influenza (strain H1N1) average duration of the exposed period range from lows of less than one day to 4 days and the average duration of the infectious period from 1.2 to 8 days [20, 32, 59]. We assume a longer exposed period to account for reporting delay. With waning immunity, individuals can be reinfected the following year. The seven years of weekly influenza data from Texas have an average total of 76,000 weekly influenza cases annually. Often many cases are not reported and a proportion of the population is vaccinated. We assume a reduced population size of 300,000 with no deaths from influenza.

### 5.2 Point Estimation of Parameters

Five parameters are estimated, *a*_*i*_, *i* = 1, 2, 3, for the periodic transmission rate 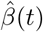, the mean reversion rate *r*_*p*_, and the diffusion coefficient *σ*_*p*_ for each of the four SEIR SDE models, *p* = 1*/*2, 2*/*3, 5*/*6, 1. Parameters are estimated with least squares fits. Again we apply a two-step procedure to estimate the parameters. First, we apply the system of SEIR ordinary differential equations (SEIR ODEs) to estimate the parameters in 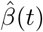 and second, we use the SDE *Y* (*t*) with the fitted mean 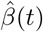 and the full SEIR SDE system to fit the parameters *r*_*p*_ and *σ*_*p*_.

We assume that 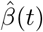 takes the form of Equation (3) and we set *Y* (*t*) equal to its mean value, 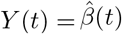, in the SEIR ODE system. Therefore, to estimate the three parameters *a*_*i*_, *i* = 1, 2, 3, in 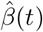, we solve the SEIR ODE system and minimize the following least squares expression:

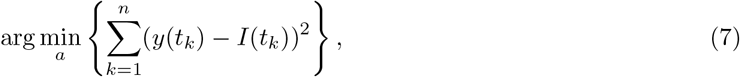

where *y*(*t*_*k*_) is the weekly influenza case data and *I*(*t*_*k*_) is the number of infectious cases at time *t*_*k*_ in the SEIR ODE system. Specifically, the parameters *a*_*i*_ in 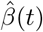 are selected such that the difference between the numerical solution *I*(*t*_*k*_) and the influenza data *y*(*t*_*k*_) are minimized. The epidemic and demographic parameters are kept fixed, *N, µ, η, γ*, and *δ*. Several parameter combinations in the SEIR model are tested within a realistic range, *η* ∈ {2, 3, 4}, *γ* ∈ {52, 104}, and *δ* ∈ {52, 104}, until the best fit is obtained for the three parameters *a*_*i*_, *i* = 1, 2, 3 (*η* = 4, *γ* = 104, and *δ* = 52). With our assumptions, we expect the ODE *I*(*t*) solution to fit the mean of the influenza data. Since the initial values of the four stages in the SEIR ODE model are unknown, we set the initial value of *I* equal to the initial value of the influenza data (519) and the initial value of *E* equal to twice this value (519× 2) because the average duration of the *E* period is twice that of *I*. Other initial values are set equal to *S* = *N*− *E* −*I* and *R* = 0. The SEIR ODE model is run for 11 years, until convergence of the SEIR model to a periodic solution. Estimates of the parameters *a*_*i*_, *i* = 1, 2, 3, in the transmission rate are chosen to give the best fit (smallest RMSE) of the data to *I*(*t*_*k*_) during the last seven years, 5-11.

The same parameter estimation method for *r*_*p*_ and *σ*_*p*_ in Section 4.1 cannot be applied to the SEIR SDE system because *I*(*t*) rather than *Y* (*t*) is being fit to the data. We test a range of *r*_*p*_ and *σ*_*p*_ values in the SEIR SDE system with a goal to minimize a weighted least squares error between the infectious cases *I*(*t*) and the seasonal influenza data. In particular, we divide the annual seasonal influenza data into five regions and denote the proportion of the data in each region as *P*_*i*_, *i* = 1, 2, 3, 4, 5 (Figure 5). The value *P*_1_ = 0.0247 is the proportion of the data *>* 5000, *P*_2_ = 0.25 is the proportion 1860 *<*data≤ 5000, *P*_3_ = 0.25 is the proportion 990 *<*data≤ 1860, *P*_4_ = 0.25 is the proportion 468 *<*data≤ 990, and *P*_5_ = 0.2253 is the proportion where data≤ 468. To ensure each proportion is equally represented, they are weighted appropriately so that all of them satisfy *w*_*i*_*P*_*i*_ = 0.25. The top proportion *P*_1_ is especially important to include so that the peak values of influenza are represented. We simulate 5000 sample paths of the SEIR SDE system for eleven years and use the last seven years, 5-11, to test a range of values for *r*_*p*_ and *σ*_*p*_, *p* = 1*/*2, 2*/*3, 5*/*6, 1. We count the total weekly influenza cases *I*(*t*) per year for each sample path that lie in one of the five regions and denote the proportion as *SPD*_*i*_, *i* = 1, 2, 3, 4, 5 (a function of the parameters *r*_*p*_ and *σ*_*p*_). The least squares values *r*_*p*_ and *σ*_*p*_ are those that minimize the following weighted least squares expression:

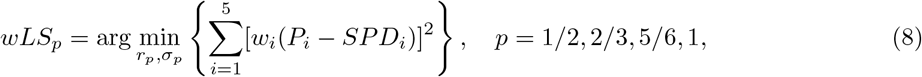

where the weights *w*_1_ = 10.11, *w*_2_ = 1 = *w*_3_ = *w*_4_, and *w*_5_ = 1.11 satisfy *w*_*i*_*P*_*i*_ = 0.25. For reference purposes, we call this method the Sample Proportion Weighted Least Squares (SPWLS). Putting the largest weight on the peak region (*SPD*_1_) ensures that sample paths reach the peak but also limits the number of values in that region, especially in the case of the large volatility with *p* = 1.

**Figure 5:**
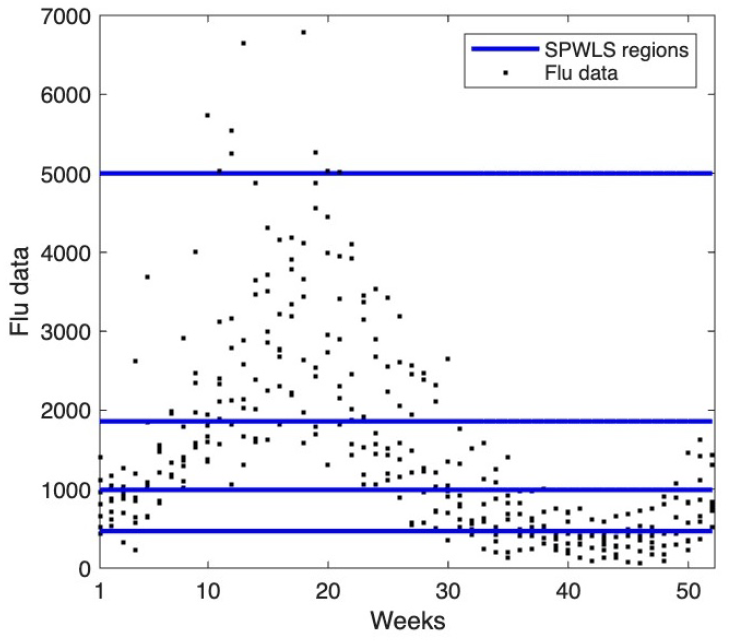
The five regions *P*_*i*_, *i* = 1, 2, 3, 4, 5, that divide the annual seasonal influenza data are identified by the blue horizontal lines separating them. Number of weeks from 1 to 52 on the *x* −axis are measured from Week 40, the beginning of seasonal influenza data. Week 13 on the *x* −axis is the first week of January.

### 5.3 Results

The estimated parameter values for the transmission rate *Y* (*t*) and the RMSEs are given in Table 3, that is, the parameters *a*_*i*_ for the mean periodic transmission rate 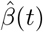 and the parameters for the reversion rate *r*_*p*_ and the volatility parameter *σ*_*p*_. The corresponding solutions 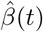, *I*(*t*), and *E*(*t*) from the SEIR ODE are annual periodic functions (Figure 10 in Appendix B). The basic reproduction number for the SEIR ODE system with the periodic transmission rate 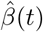 can be estimated using methods from [74]: R_0_ = 1.158. This estimate is close to values of ℛ_0_ in other seasonal influenza models [11].

**Table 3:** Estimates of the five parameter values for the transmission rate *Y* (*t*) in the SEIR SDE epidemic model and their RMSEs. Parameters for *r*_*p*_ and *σ*_*p*_ are to nearest 0.05 unit.

| *a*<sub>1</sub> | *a*<sub>2</sub> | *a*<sub>3</sub> | RMSE |
|---|---|---|---|
| 130.8 | 0.8680 | 0.2711 | 769.3 |
| *p* | *r*<sub>*p*</sub> | *σ*<sub>*p*</sub> | RMSE |
| 1/2 | 7.25 | 4.15 | 3.506 × 10<sup>−2</sup> |
| 2/3 | 7.70 | 2.00 | 3.298 × 10<sup>−2</sup> |
| 5/6 | 8.35 | 0.95 | 3.210 × 10<sup>−2</sup> |
| 1 | 9.75 | 0.45 | 2.926 × 10<sup>−2</sup> |

The parameters in Table 3 show the same trend as *p* increases from 1/2 to 1 as in Table 1; the mean reversion rate *r*_*p*_ increases while *σ*_*p*_ decreases. One difference in the two fits is that best fit of the data to the SEIR SDE models is the case *p* = 1 and the worst fit is for the case *p* = 1*/*2, the reverse of the fit of the SDEs directly to the data (Table 1). The sample proportion data from the SPWLS method in Table 4 reveal a few differences between the four SEIR SDEs. The sample proportions *SPD*_*i*_, *i* = 1, 2, 3, 4, are all less than the original data *P*_*i*_, *i* = 1, 2, 3, 4, for cases *p* = 1*/*2, 2*/*3, 5*/*6, with the greatest proportion allotted to region *SPD*_5_. Whereas for the case *p* = 1, greater proportions are allotted to two regions *SPDI*_3_ and *SPD*_4_ and the proportion *SPD*_5_ is closer to the data (Tables 3-4). However, the small differences between the values of the RMSEs in Table 3 indicate small differences between the fits.

**Table 4:**
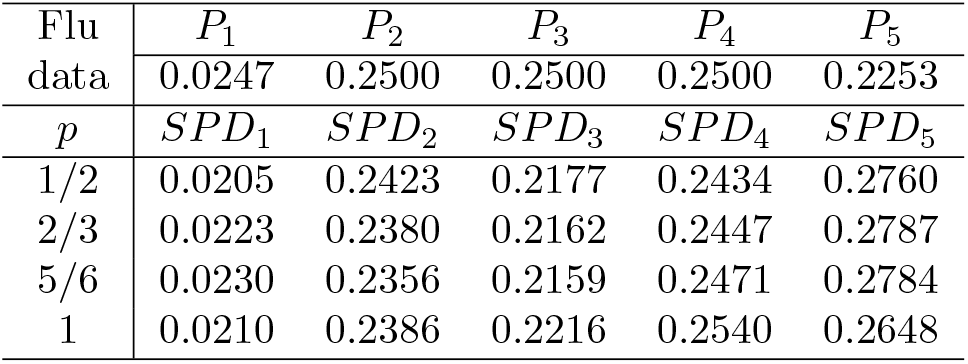
The proportions *P*_*i*_ from the influenza data are compared to the sample proportions *SPD*_*i*_, *i* = 1, 2, 3, 4, 5 from the SPWLS method obtained from 5000 sample paths for each of the SEIR models, *p* = 1*/*2, 2*/*3, 5*/*6, 1.

| Flu data | *P*<sub>1</sub> | *P*<sub>2</sub> | *P*<sub>3</sub> | *P*<sub>4</sub> | *P*<sub>5</sub> |
|  | 0.0247 | 0.2500 | 0.2500 | 0.2500 | 0.2253 |
| *p* | *SPD*<sub>1</sub> | *SPD*<sub>2</sub> | *SPD*<sub>3</sub> | *SPD*<sub>4</sub> | *SPD*<sub>5</sub> |
|---|---|---|---|---|---|
| 1/2 | 0.0205 | 0.2423 | 0.2177 | 0.2434 | 0.2760 |
| 2/3 | 0.0223 | 0.2380 | 0.2162 | 0.2447 | 0.2787 |
| 5/6 | 0.0230 | 0.2356 | 0.2159 | 0.2471 | 0.2784 |
| 1 | 0.0210 | 0.2386 | 0.2216 | 0.2540 | 0.2648 |

The fit of the periodic SEIR ODE solution *I*(*t*) (denoted ‘ODE *I*(*t*)’) is compared directly to the mean of the influenza data and to the periodic function *β*(*t*) from Equation (1). Shown in the left panel of Figure 6 are graphs of the two periodic functions and the mean of the data and in the right panel the periodic mean transmission rate 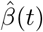 and the solution ODE *I*(*t*). Although the ODE *I*(*t*) differs slightly from *β*(*t*) with larger maxima and minima than *β*(*t*), the two periodic functions are aligned well with the mean of the seasonal influenza data. Another similarity between the two functions is that they agree in their average total number of weekly influenza cases per year, i.e.,

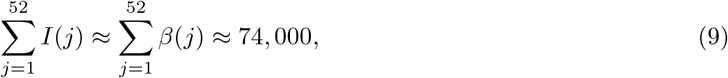

**Figure 6:**
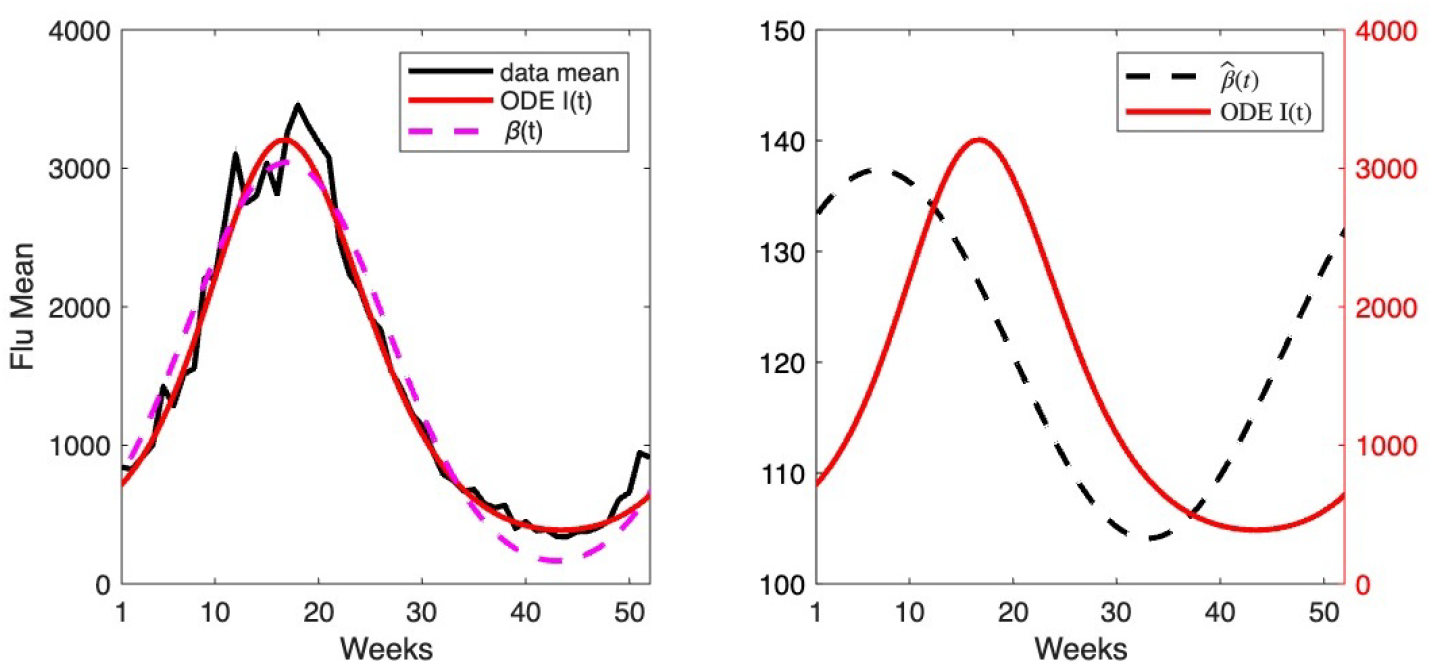
Left panel: Graphs of the average weekly number of reported influenza cases (solid black curve), the periodic mean function *β*(*t*) in Equation (3) (dashed magenta curve) and the solution ODE *I*(*t*) of the SEIR model (solid red curve). Right panel: Graphs of the periodic transmission rate 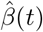 (dashed black curve) and the periodic solution ODE *I*(*t*) (solid red curve); units on y-axis on the left are for 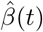 (per year) and units on the right are for ODE *I*(*t*) (weekly flu cases). Week 1 on the x-axis is equal to Week 40. Therefore, Week 12 on the x-axis is the last week of December and Week 13 is the first week of January.

The times of occurrence of the highest peaks and minima for the two periodic functions, *β*(*t*) and ODE *I*(*t*), are also in agreement in Figure 6. For the ODE *I*(*t*), the time of the highest peak (measured from January 1st) is week 4.7 (≈first week of February), whereas for *β*(*t*) the time of the highest peak is week 5. These values are in agreement with the influenza data, where the average time for the peak is week 5. The times of lowest number of influenza cases vary in the data. If in each year, we take the minimum of the average of three consecutive data values and the average over seven years, we obtain week 28 (≈mid July), the off-season for influenza. The two functions, ODE *I*(*t*) and *β*(*t*), have their lowest values about 3 weeks later, in weeks 31.4 and 31, respectively. An interesting phenomenon seen in the right panel of Figure 6 is that the times of the maxima and minima of the periodic transmission rate 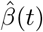 precede those of the ODE *I*(*t*) and differ by about 10 weeks. This difference in timing is seen in other deterministic models with a periodic transmission rate [43]. For the mean of the transmission rate, 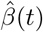, the peak appears about week 47 (≈end of November) and the minimum about week 21 (≈end of May), whereas for the ODE *I*(*t*) the time of the peak is week 4.7 (≈first week of February) and the minimum about week 31.4 (≈ first week in August).

Next, we graph sample paths for the infectious cases *I*(*t*) and the transmission rate *Y* (*t*) associated with the four SEIR SDE models in Figure 7. In the four panels of the first row are the sample paths for SDE *I*(*t*) and the periodic ODE *I*(*t*) solution. In the second row are the corresponding sample paths of the transmission rate SDE *Y* (*t*) and the periodic function 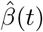. The sample paths for the influenza cases *I*(*t*) show that many of the major peaks each year are not attained. There is less variability of the infectious cases about the mean than in the first model (Figure 2) as the transmission rate *Y* (*t*) is modulated by the dynamics of susceptible and infectious stages through the term *Y* (*t*)*S*(*t*)*I*(*t*)*/N*. Different phenomena are modeled in the SDEs for *X*(*t*) (number of cases) versus those for *Y* (*t*) (transmission rate) and also the objectives of the two fitting methods differ. The maximum likelihood method applies the point data as a function of time, whereas the SPWLS applies the proportional data. This latter method needs further adjustment to ensure peaks are reached.

**Figure 7:**
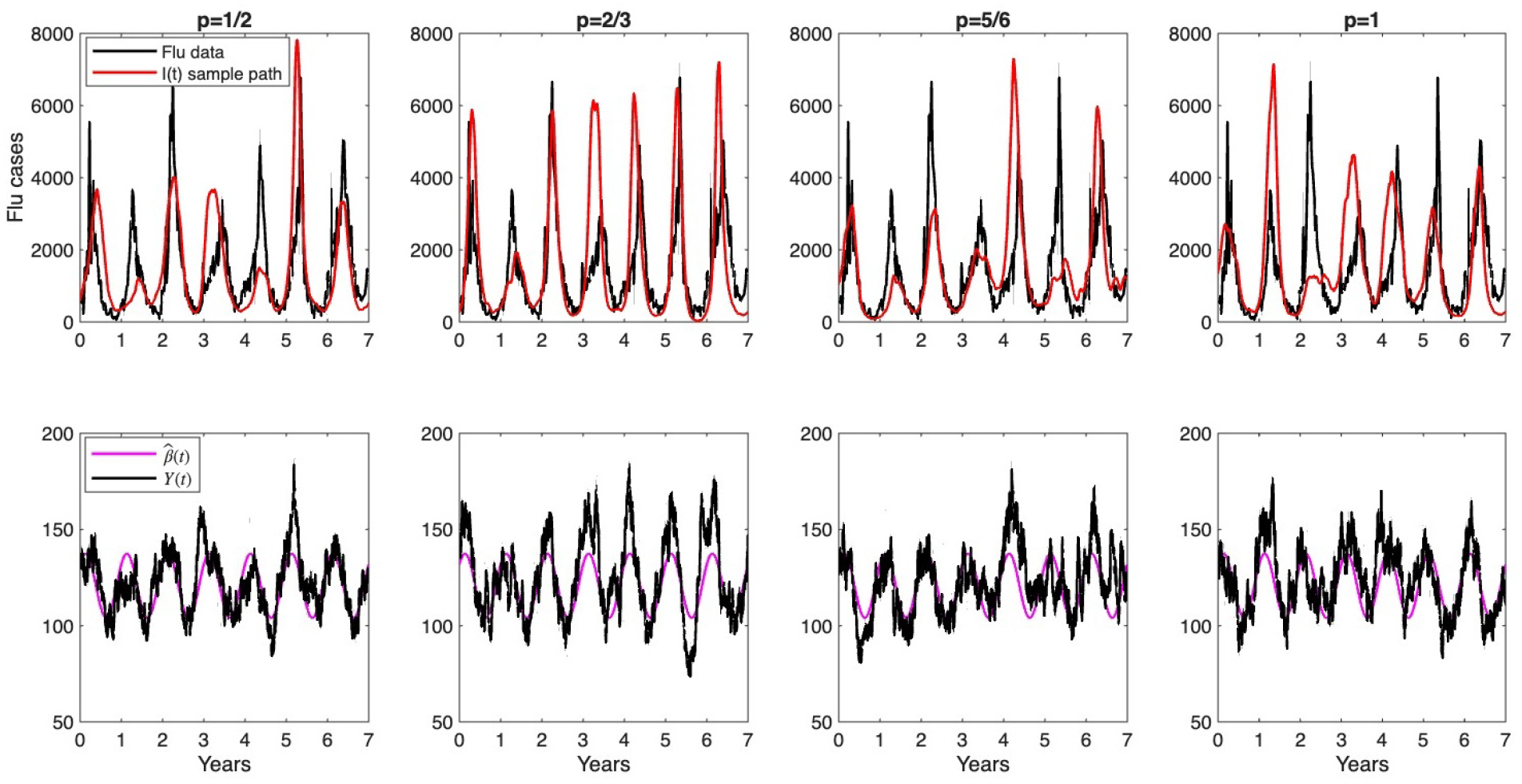
Top row: One sample path of the SDE *I*(*t*) from each of the four SEIR SDE models, *p* = 1*/*2, 2*/*3, 5*/*6, 1, (red curve) is plotted with the influenza data (black curve). Bottom row: One sample path of the corresponding seasonal mean-reverting transmission rate, SDE *Y* (*t*), for each *p* = 1*/*2, 3*/*4, 5*/*6, 1 (black curve) is plotted with the mean periodic transmission rate 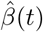 (magenta curve).

When we examine the time of the major peaks in Figure 5 we find variability in the weeks that they occur over the seven years. For example, in case *p* = 2*/*3, the five major peaks occur in weeks 4, 1.9, 1.2, 0.3, 3.1, 3.7 (early January to February). The maximum peak in the other cases *p* = 1*/*2, 5*/*6, 1 occur in weeks 1.8 (January, year 6), week 6.7 (February, year 5), and week 6.4 (February, year 2), respectively. The variability in peaks is somewhat consistent with the times of the highest peak in the seven years of influenza data, weeks 52, 2, 1, 11, 7, 6, and 8, but the sample paths are only a small representation of the peaks. Long-term seasonal influenza data for the United States show peak number of cases occur most often in February followed by December [18] (Appendix B). The shift in the peaks in the influenza data from December/January to February/March may be due to a shift in the dominant viral type or subtype (A H1N1/H3N2 or B).

As a final example, we compare sample paths of *I*(*t*) for the mean-reverting SEIR SDE model and the mean-reverting SDE model from Section 4. In the left panel of Figure 8 is graphed the influenza data. The center panel is a sample path of *I*(*t*) for the mean-reverting SEIR SDE and in the right panel for the mean-reverting SDE *X*(*t*). One obvious difference between these two models is the variability. Sample paths of *X*(*t*) are highly variable and those of the SEIR SDE *I*(*t*) are smooth. The variability in the influenza data lies between these two extremes. Although, the SEIR SDE *I*(*t*) does not always reach the peaks, the variability in the timing of the peaks, the range of the peaks, and the stages in the structured SEIR model provide a realistic and meaningful setting for interpretation of the epidemiological results over many seasons.

**Figure 8:**
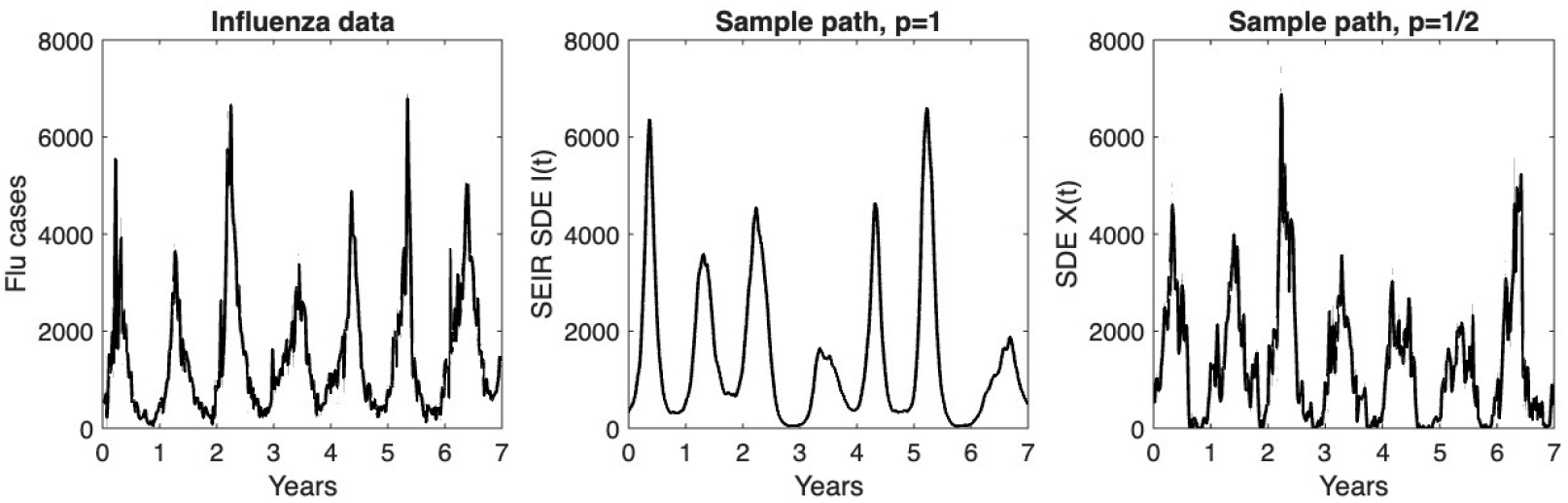
Left panel: Graph of the influenza data; Center panel: Sample path of *I*(*t*), the best fit for the mean-reverting SEIR SDE model (*p* = 1); Right panel: Sample path of the best fit for the mean-reverting SDE *X*(*t*) (*p* = 1*/*2).

## 6 Discussion

The novel periodic mean-reverting SDEs in Equation (1) provide a realistic framework for capturing seasonal variation in a variety of environmental and biological systems. Although such models are common in financial and economic applications, e.g. [28, 29, 61, 53], they have not been investigated in an epidemic setting.

This investigation focuses on periodic mean-reverting SDEs that incorporate state-dependent diffusion which effectively accounts for the complexities of seasonal processes and ensures solutions are nonnegative. Seasonal influenza data are used to estimate model parameters in two different settings. First, the mean-reverting processes in Equation (1) are fit directly to the seasonal influenza data, and second, they are used as a model for the transmission rate, where the infectious cases *I* in an SEIR epidemic model are fit to the influenza data.

In the first setting, the Euler–Maruyama approximation provides a stable computationally efficient likelihood-based framework for estimation of parameters when exact transition densities are unavailable. For the more complex SEIR structured models, a sample proportion weighted least squares method (SP-WLS), fit to the proportional data, helps to ensure the peaks of the seasons are adequately represented. Because the influenza observations are available at weekly intervals, some discretizations error may nevertheless remain. Alternative approaches include quasi-maximum likelihood and continuous-discrete filtering methods, such as the extended Kalman filter, which are particularly useful for discretely observed continuous-time stochastic processes [28, 44, 55]. These methods provide potential extensions for assessing sensitivity of parameter estimates to the transition-density approximation, while the Euler–Maruyama and weighted least squares approaches adopted here offer transparent and consistent frameworks for comparing the alternative diffusion specifications.

Understanding the dynamics of the periodic mean-reverting SDE in Equation (1) and the relation to the parameter estimates are important because it directly informs how uncertainty and stability evolve in the modeled system. The diffusion term *σ*_*p*_*X*^*p*^ governs the strength of the stochastic perturbations, increasing markedly as *p* increases (Figure 4) indicating that the system becomes more sensitive to random fluctuations. This has significant implications for modeling biological and physical systems where noise plays a critical role. Higher values of *p* (close to one) may represent regimes of reduced predictability or control. Furthermore, the observed decline in the correlation between *r*_*p*_ and *σ*_*p*_ for midrange values of *p* = 2*/*3, 5*/*6 suggests a decoupling of the systems’ reversion dynamics from its stochastic volatility, allowing for more precise tuning of each component.

Through leverage of bootstrapping techniques, we constructed confidence regions for parameter estimation, improving the robustness of inference and revealing trends in parameter relations. The demonstrated ability to fit parameters of seasonal mean-reverting SDEs and to obtain confidence regions for the parameters, enhances the flexibility and applicability of these time series models.

The transmission rate in epidemiological models is one of the most common parameters to estimate but the parameters for mean reversion *r*_*p*_ and volatility *σ*_*p*_ are hard to quantify in epidemiological terms. In a COVID-19 epidemic model with a constant mean-reverting transmission, the mean-reversion rate was interpreted as a response to restrictions (e.g., social distancing and masks) imposed during the pandemic [63]. Our model was able to capture some of the variability seen in the influenza data, including the timing and variability of the peaks and the average annual seasonality (Figures 6 and 7). In addition, the SEIR model showed that the peak transmission rate can precede the peak influenza cases by several months (Figure 6, right panel); it was approximately 10 weeks from the peak of 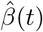 to the peak of the ODE *I*(*t*). This has implications for public health practitioners, highlighting the need for early interventions. Also, as shown in Figure 3, the parameters *r*_*p*_ and *σ*_*p*_ can be highly correlated. In the best fit models, the SEIR SDE model has a longer average time to return to the mean, 364*/r*_1_ ≈ 37.3 days (*p* = 1) versus the direct fit of the mean-reverting SDE Equation (1) to the data, which has a return time of 364*/r*_1*/*2_ ≈ 28.4 days (*p* = 1*/*2). A longer duration allows for a longer span of time above or below the mean. Model fit can be improved with more data on the impact of social behavior and climate variability on mean reversion. Also, data on the circulating viral types and subtypes (A H1N1/H3N2, B), the duration of immunity, and the vaccine coverage will improve model predictability, e.g., [8, 19, 26, 45, 73]. This foundational study of periodic mean-reverting processes serves as a basis for further investigation of more complex structured epidemic models and for application of alternative statistical methods to fit long-term data sets that capture the important features of seasonality. Insights from these models are essential for calibrating parameters, enabling better forecasts, risk assessment, and control strategies in complex stochastic environments.

Moreover, an important consideration is the potential identifiability between the deterministic seasonal component and stochastic diffusion component, since both can explain temporal variation in influenza activity. In particular, excessive flexibility in the periodic mean may absorb short-term stochastic variation, whereas a large diffusion coefficient may obscure the underlying seasonal signal. Our two-stage estimation strategy helps separate these components by first estimating the systematic periodic structure and subsequently estimating the stochastic parameters conditional on that structure. Nevertheless, uncertainty in the estimated seasonal component may propagate to the diffusion parameter estimates, motivating sensitivity and bootstrap analyses.

Overall, in public health, the ability to anticipate and respond to disease outbreaks hinges not only on recognizing the seasonal cycles that shape predictable patterns of transmission, but also on effectively incorporating stochastic models that capture uncertainty and variability in real-world dynamics. By embracing stochastic approaches, epidemiology moves beyond rigid forecasts to provide probabilistic insights that reflect the inherent randomness of disease spread, offering health authorities better tools for risk assessment, preparedness, and resource allocation. These new approaches help ensure that interventions are guided by a realistic understanding of both expected seasonal trends and the unpredictable shocks that often define epidemic behavior.

## Data Availability

Data is available Texas DSHS website. https://www.dshs.texas.gov/influenza-flu/influenza-flu-surveillance

## Acknowledgments

We thank the editor and the three anonymous reviewers for their helpful and insightful suggestions to expand the discussion on alternative stochastic models, uncertainty issues, and epidemiological interpretation. The authors gratefully acknowledge the assistance of EJ Allen for suggesting the method for estimating the RMSE in Equation (6).

## Author Contributions

FM: data collection and machine learning method; FM & LJSA: conceptualization, formal analysis, methodology, computer codes, numerical simulations, and writing.

## Data Availability

The datasets are available at the Texas Department of State Health Services recorded in the Texas Influenza Surveillance Activity Reports for years 2012-2019 on the website: https://www.dshs.texas.gov/influenza-flu-provider-information/influenza-flu-surveillance.

## Appendix A MissForest Imputation

Figure 9 is an illustration of the imputation of missing weekly influenza cases for the MissForest algorithm. On the right side of Figure 9, white boxes labeled ‘NAN’ indicate missing values across different years and weeks. The left side displays the same dataset after imputation, where the previously missing values are filled in and highlighted in light blue. This visual comparison demonstrates how the MissForest algorithm effectively reconstructs a complete dataset suitable for time series analysis.

**Figure 9:**
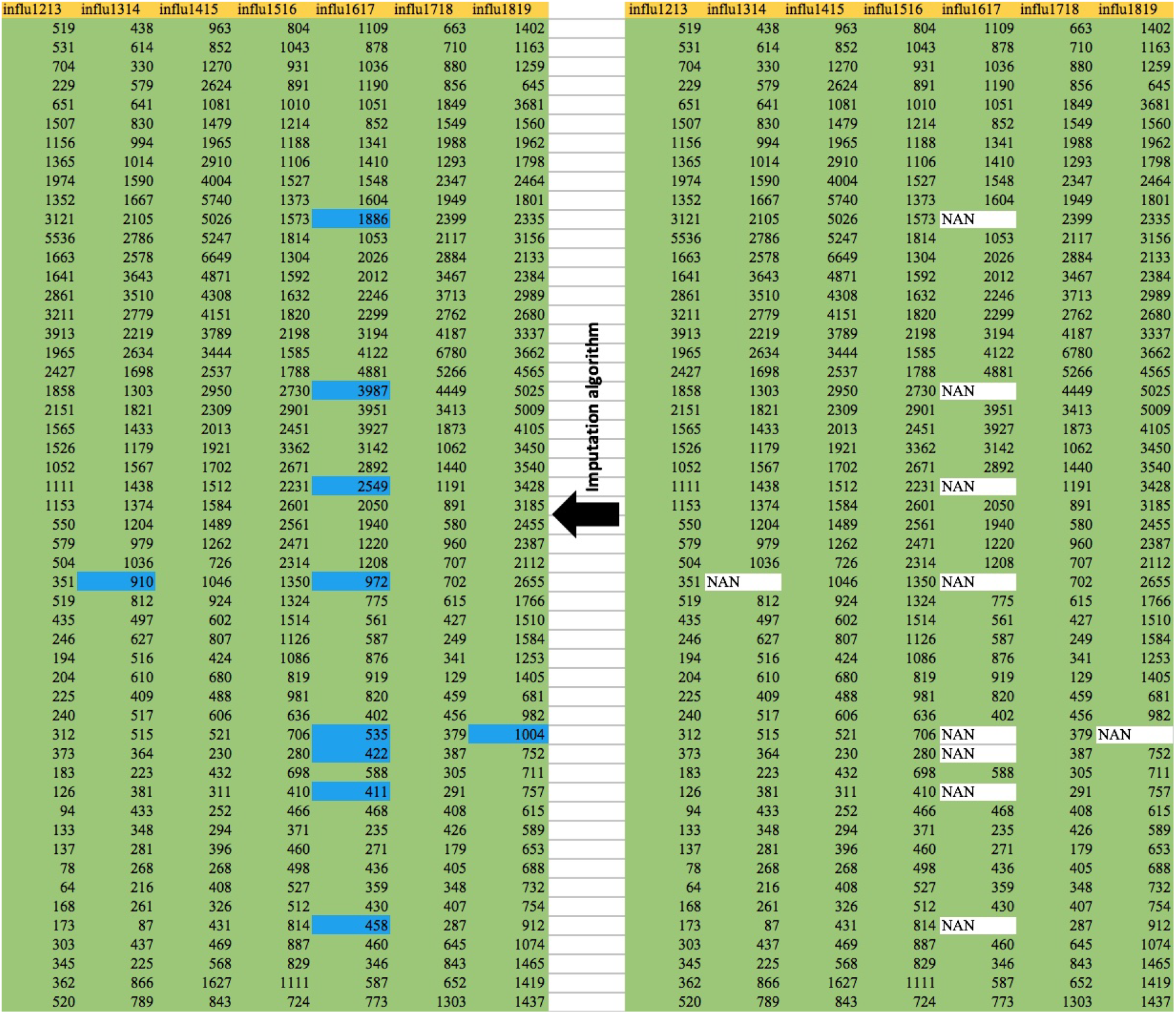
On the right, NAN white boxes represent missing data points (weekly cases) during weeks 1-52 during the year. On the left, imputed data with light blue boxes are presented after applying the MissForest algorithm.

## Appendix B SEIR Supplementary Figures and Data

In Figure 10 are graphs of the periodic mean transmission rate 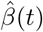, its mean= 120.7, and the periodic solutions for the infectious stage *I*(*t*) and exposed stage *E*(*t*) when the transmission rate in the SEIR ODE model equals 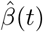.

**Figure 10:**
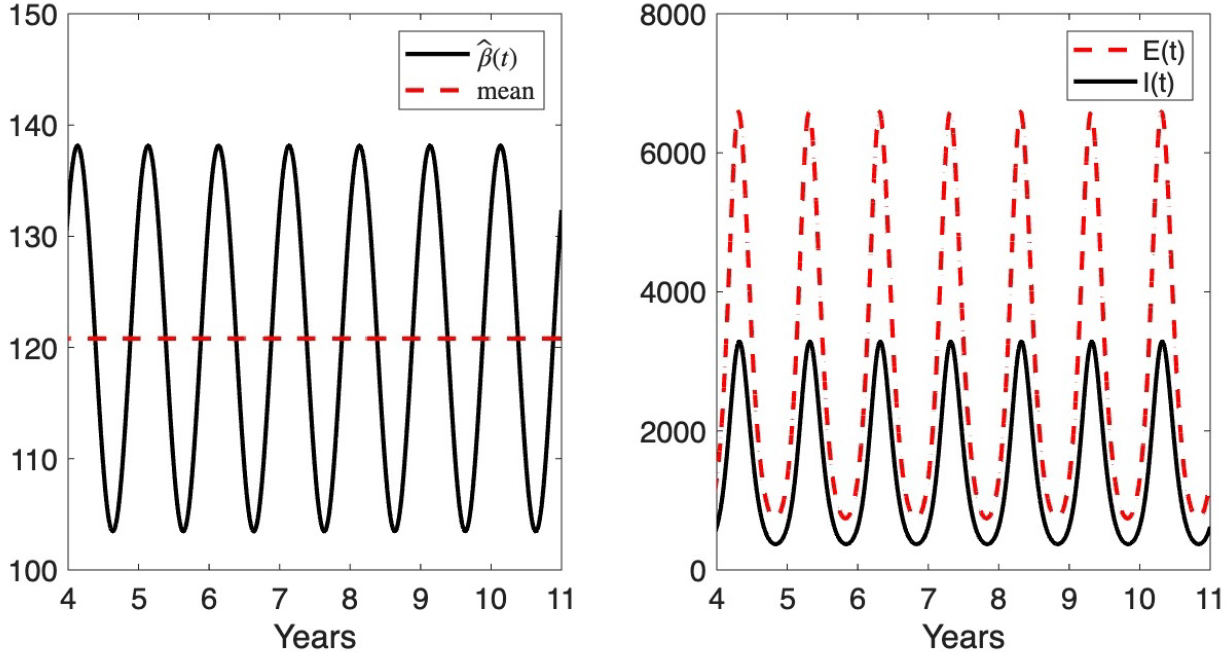
Graphs of the periodic transmission rate 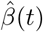 with mean = 120.7 and the periodic solutions of the infectious stage *I*(*t*) and exposed stage *E*(*t*) for the SEIR ODE model over 7 years, years 5-11.. Parameters estimates *a*_*i*_, *i* = 1, 2, 3 for 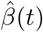 are given in Table 3.

Data compiled from the Centers for Disease Control and Prevention [18] record the months with greatest seasonal influenza activity in the United States over a period of 42 years, 1982-83 through 2024-2025, which are summarized in Figure 11.

**Figure 11:**
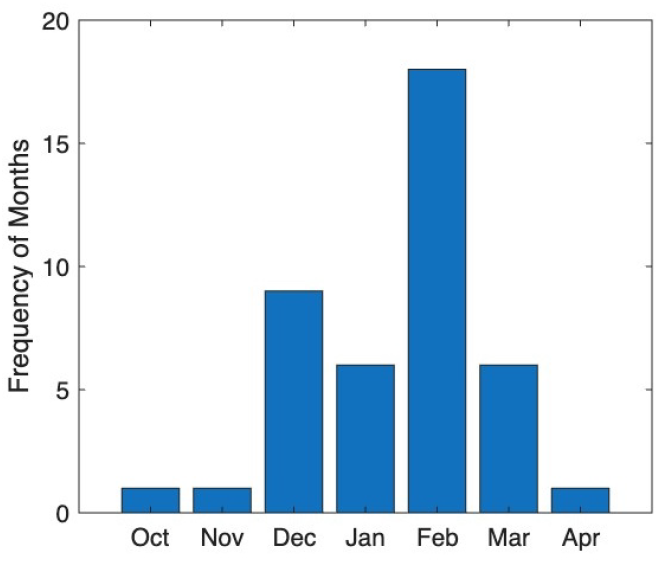
Months with greatest flu activity in U.S.A. during the years from 1982-83 to 2024-25 [18].

## References

[1] H. Akima. A new method of interpolation and smooth curve fitting based on local procedures. Journal of the ACM (JACM), 17(4):589–602, 1970.

[2] S. T. Ali and B. J. Cowling. Influenza virus: tracking, predicting, and forecasting. Annual Review of Public Health, 42(1):43–57, 2021.

[3] E. Allen. Modeling with Itô Stochastic Differential Equations, volume 22. Springer Science & Business Media, Dordrecht, 2007.

[4] E. Allen. Environmental variability and mean-reverting processes. Discrete Contin. Dyn. Syst. Ser. B, 21(7):2073–2089, 2016.

[5] E. J. Allen, L. J. S. Allen, and H. L. Smith. On real-valued SDE and nonnegative-valued SDE population models with demographic variability. Journal of Mathematical Biology, 81(2):487–515, 2020.

[6] S. Altizer, A. Dobson, P. Hosseini, P. Hudson, M. Pascual, and P. Rohani. Seasonality and the dynamics of infectious diseases. Ecology Letters, 9(4):467–484, 2006.

[7] C. Andreu-Vilarroig, G. González-Parra, and R.-J. Villanueva. Mathematical modeling of influenza dynamics: Integrating seasonality and gradual waning immunity. Bulletin of Mathematical Biology, 87(6):75, 2025.

[8] A. Andronico, J. Paireau, and S. Cauchemez. Integrating information from historical data into mechanistic models for influenza forecasting. PLOS Computational Biology, 20(10):e1012523, 2024.

[9] N. T. Bailey. The Mathematical Theory of Infectious Diseases and its Applications. Charles Griffin and Company, 1975.

[10] D. Balcan, H. Hu, B. Goncalves, P. Bajardi, C. Poletto, J. J. Ramasco, D. Paolotti, N. Perra, M. Tizzoni, W. Van den Broeck, et al. Seasonal transmission potential and activity peaks of the new influenza A (H1N1): a Monte Carlo likelihood analysis based on human mobility. BMC Medicine, 7(1):45, 2009.

[11] M. Biggerstaff, S. Cauchemez, C. Reed, M. Gambhir, and L. Finelli. Estimates of the reproduction number for seasonal, pandemic, and zoonotic influenza: a systematic review of the literature. BMC Infectious Diseases, 14(1):1–20, 2014.

[12] L. Billings and E. Forgoston. Seasonal forcing in stochastic epidemiology models. Ricerche di Matematica, 67(1):27–47, 2018.

[13] F. Black and M. Scholes. The pricing of options and corporate liabilities. Journal of political economy, 81(3):637–654, 1973.

[14] G. E. Box, G. M. Jenkins, G. C. Reinsel, and G. M. Ljung. Time Series Analysis: Forecasting and Control. John Wiley & Sons, 2015.

[15] S. Brooks, A. Gelman, G. Jones, and X.-L. Meng. Handbook of Markov Chain Monte Carlo. CRC press, 2011.

[16] Y. Cai, J. Jiao, Z. Gui, Y. Liu, and W. Wang. Environmental variability in a stochastic epidemic model. Applied Mathematics and Computation, 329:210–226, 2018.

[17] A. C. Cameron and P. K. Trivedi. Regression Analysis of Count Data. Number 53. Cambridge University Press, 2013.

[18] Centers for Disease Control and Prevention. Influenza (Flu), Flu Season. https://www.cdc.gov/flu/about/season.html, 2025. Accessed: 2026-08-24.

[19] K. Charland, D. Buckeridge, J. Sturtevant, F. Melton, B. Reis, K. Mandl, and J. Brownstein. Effect of environmental factors on the spatio-temporal patterns of influenza spread. Epidemiology & Infection, 137(10):1377–1387, 2009.

[20] A. Cori, A.-J. Valleron, F. Carrat, G. S. Tomba, G. Thomas, and P.-Y. Boëlle. Estimating influenza latency and infectious period durations using viral excretion data. Epidemics, 4(3):132–138, 2012.

[21] J. C. Cox, J. E. Ingersoll Jr, and S. A. Ross. A theory of the term structure of interest rates. Econometrica, 53(2):385–407, 1985.

[22] W. Dai. Nonexistence of positive solutions to n-th order equations in Rn. Bulletin des Sciences Mathématiques, 174:103072, 2022.

[23] K. Dietz. The incidence of infectious diseases under the influence of seasonal fluctuations. In Mathematical Models in Medicine: Workshop, Mainz, March 1976, pages 1–15. Springer, 1976.

[24] V. Dukic, H. F. Lopes, and N. G. Polson. Tracking epidemics with google flu trends data and a state-space seir model. Journal of the American Statistical Association, 107(500):1410–1426, 2012.

[25] J. Durbin and S. J. Koopman. Time Series Analysis by State Space Methods. Oxford University Press (UK), 2012.

[26] S. Edlund, J. Kaufman, J. Lessler, J. Douglas, M. Bromberg, Z. Kaufman, R. Bassal, G. Chodick, R. Marom, V. Shalev, et al. Comparing three basic models for seasonal influenza. Epidemics, 3(3–4):135–142, 2011.

[27] P. Esling and C. Agon. Time-series data mining. ACM Computing Surveys (CSUR), 45(1):1–34, 2012.

[28] C. Ewald and Y. Zou. Analytic formulas for futures and options for a linear quadratic jump diffusion model with seasonal stochastic volatility and convenience yield: Do fish jump? European Journal of Operational Research, 294(2):801–815, 2021.

[29] C.-O. Ewald and R. Ouyang. An analysis of the fish pool market in the context of seasonality and stochastic convenience yield. Marine Resource Economics, 32(4):431–449, 2017.

[30] W. F. Fagan, F. McBride, and L. Koralov. Reinforced diffusions as models of memory-mediated animal movement. Journal of the Royal Society Interface, 20(200):20220700, 2023.

[31] D. N. Fisman. Seasonality of infectious diseases. Annual Review of Public Health, 28(1):127–143, 2007.

[32] C. Fraser, C. A. Donnelly, S. Cauchemez, W. P. Hanage, M. D. Van Kerkhove, T. D. Hollingsworth, J. Griffin, R. F. Baggaley, H. E. Jenkins, E. J. Lyons, et al. Pandemic potential of a strain of influenza A (H1N1): early findings. Science, 324(5934):1557–1561, 2009.

[33] A. R. Gallant and G. Tauchen. The relative efficiency of method of moments estimators. Journal of Econometrics, 92(1):149–172, 1999.

[34] P. Gloaguen, M.-P. Etienne, and S. Le Corff. Stochastic differential equation based on a multimodal potential to model movement data in ecology. Journal of the Royal Statistical Society Series C: Applied Statistics, 67(3):599–619, 2018.

[35] S. He, S. Tang, Y. Cai, W. Wang, and L. Rong. A stochastic epidemic model coupled with seasonal air pollution: analysis and data fitting. Stochastic Environmental Research and Risk Assessment, 34:2245–2257, 2020.

[36] L. Held, M. Höhle, and M. Hofmann. A statistical framework for the analysis of multivariate infectious disease surveillance counts. Statistical Modelling, 5(3):187–199, 2005.

[37] L. Held and M. Paul. Modeling seasonality in space-time infectious disease surveillance data. Bio-metrical Journal, 54(6):824–843, 2012.

[38] U. Helfenstein. Box-Jenkins modelling of some viral infectious diseases. Statistics in Medicine, 5(1):37–47, 1986.

[39] D. J. Higham. An algorithmic introduction to numerical simulation of stochastic differential equations. SIAM Review, 43(3):525–546, 2001.

[40] E. M. Hill, S. Petrou, S. de Lusignan, I. Yonova, and M. J. Keeling. Seasonal influenza: Modelling approaches to capture immunity propagation. PLoS Computational Biology, 15(10):e1007096, 2019.

[41] A. S. Hurn, K. A. Lindsay, and V. L. Martin. On the efficacy of simulated maximum likelihood for estimating the parameters of stochastic differential equations. Journal of Time Series Analysis, 24(1):45–63, 2003.

[42] S. M. Iacus. Simulation and Inference for Stochastic Differential Equations: with R Examples, volume 486. Springer-Verlag, New York, 2008.

[43] M. J. Keeling and P. Rohani. Modeling Infectious Diseases in Humans and Animals. Princeton University Press, 2008.

[44] N. R. Kristensen, H. Madsen, and S. B. Jørgensen. Parameter estimation in stochastic grey-box models. Automatica, 40(2):225–237, 2004.

[45] L. Li, Y. Jiang, and B. Huang. Long-term prediction for temporal propagation of seasonal influenza using transformer-based model. Journal of Biomedical Informatics, 122:103894, 2021.

[46] R. J. Little and D. B. Rubin. Statistical Analysis with Missing Data. John Wiley & Sons, 2019.

[47] E. Lofgren, N. H. Fefferman, Y. N. Naumov, J. Gorski, and E. N. Naumova. Influenza seasonality: underlying causes and modeling theories. Journal of Virology, 81(11):5429–5436, 2007.

[48] W.-Y. Loh, Q. Zhang, W. Zhang, and P. Zhou. Missing data, imputation and regression trees. Statistica Sinica, 30(4):1697–1722, 2020.

[49] K. Mamis and M. Farazmand. Stochastic compartmental models of the COVID-19 pandemic must have temporally correlated uncertainties. Proceedings of the Royal Society, A, 479:2022056, 2023.

[50] P. McCullagh. Generalized Linear Models. Routledge, 2019.

[51] R. Merton. Theory of rational option pricing. Bell Journal of Economics and Management Science, 4:141–183, 1973.

[52] T. Michelot, R. Glennie, C. Harris, and L. Thomas. Varying-coefficient stochastic differential equations with applications in ecology. Journal of Agricultural, Biological and Environmental Statistics, 26:446–463, 2021.

[53] M. Moreno, A. Novales, and F. Platania. Long-term swings and seasonality in energy markets. European Journal of Operational Research, 279(3):1011–1023, 2019.

[54] F. Mostafa and L. J. S. Allen. Periodic mean-reverting stochastic differential equations and parameter estimations for seasonal data. Stochastic Models, pages 1–32, 2025.

[55] J. N. Nielsen and H. Madsen. Applying the EKF to stochastic differential equations with level effects. Automatica, 37(1):107–112, 2001.

[56] K. F. Nipa, S. R. J. Jang, and L. J. S. Allen. The effect of demographic and environmental variability on disease outbreak for a dengue model with a seasonally varying vector population. Mathematical Biosciences, 331:108516, 2021.

[57] D. Osthus, K. S. Hickmann, P. C. Caragea, D. Higdon, and S. Y. Del Valle. Forecasting seasonal influenza with a state-space SIR model. The Annals of Applied Statistics, 11(1):202, 2017.

[58] U. Picchini, S. Ditlevsen, and A. De Gaetano. Maximum likelihood estimation of a time-inhomogeneous stochastic differential model of glucose dynamics. Mathematical Medicine and Biology: a Journal of the IMA, 25(2):141–155, 2008.

[59] B. Pourbohloul, A. Ahued, B. Davoudi, R. Meza, L. A. Meyers, D. M. Skowronski, I. Villasenor, F. Galvan, P. Cravioto, D. J. Earn, et al. Initial human transmission dynamics of the pandemic (H1N1) 2009 virus in North America. Influenza and Other Respiratory Viruses, 3(5):215–222, 2009.

[60] A. C. Rencher and W. F. Christensen. Methods of Multivariate Analysis, Wiley Series in Probability and Statistics, volume 709. John Wiley & Sons, New Jersey, USA, 2012.

[61] M. Richter and C. Sørensen. Stochastic volatility and seasonality in commodity futures and options: The case of soybeans. Available SSRN:, 2002.

[62] S. V. Scarpino and G. Petri. On the predictability of infectious disease outbreaks. Nature Communications, 10(1):898, 2019.

[63] N. Shakiba, C. J. Edholm, B. O. Emerenini, A. L. Murillo, A. Peace, O. Saucedo, X. Wang, and L. J. S. Allen. Effects of environmental variability on superspreading transmission events in stochastic epidemic models. Infectious Disease Modelling, 6:560–583, 2021.

[64] J. Shaman and A. Karspeck. Forecasting seasonal outbreaks of influenza. Proceedings of the National Academy of Sciences, 109(50):20425–20430, 2012.

[65] K. P. Sharp. Stochastic differential equations in finance. Applied Mathematics and Computation, 37(2):131–148, 1990.

[66] R. H. Shumway and D. S. Stoffer. Time Series Analysis and its Applications: with R examples. Springer, 2006.

[67] D. J. Stekhoven and P. Bühlmann. Missforest—non-parametric missing value imputation for mixedtype data. Bioinformatics, 28(1):112–118, 2012.

[68] L. Stone, R. Olinky, and A. Huppert. Seasonal dynamics of recurrent epidemics. Nature, 446(7135):533–536, 2007.

[69] T. Su, Q. Yang, X. Zhang, and D. Jiang. Stationary distribution, extinction and probability density function of a stochastic SEIV epidemic model with general incidence and Ornstein–Uhlenbeck process. Physica A: Statistical Mechanics and its Applications, 615:128605, 2023.

[70] M. Templ. Nonlinear methods. In Visualization and Imputation of Missing Values: With Applications in R, pages 273–323. Springer, 2023.

[71] Texas-DSHS. Influenza (Flu) — Texas DSHS — dshs.texas.gov. https://www.dshs.texas.gov/influenza-flu-provider-information/influenza-flu-surveillance, n.d. [Accessed 01-01-2021].

[72] T. Tian, Y. Zhou, Y. Wu, and X. Ge. Estimation of parameters in mean-reverting stochastic systems. Mathematical Problems in Engineering, 2014:317059, 2014.

[73] J. Truscott, C. Fraser, S. Cauchemez, A. Meeyai, W. Hinsley, C. A. Donnelly, A. Ghani, and N. Ferguson. Essential epidemiological mechanisms underpinning the transmission dynamics of seasonal influenza. Journal of The Royal Society Interface, 9(67):304–312, 2012.

[74] W. Wang and X.-Q. Zhao. Threshold dynamics for compartmental epidemic models in periodic environments. Journal of Dynamics and Differential Equations, 20(3):699–717, 2008.

[75] M. West and J. Harrison. Bayesian Forecasting and Dynamic Models. Springer, 1997.

[76] W. Yang, M. Lipsitch, and J. Shaman. Inference of seasonal and pandemic influenza transmission dynamics. Proceedings of the National Academy of Sciences, 112(9):2723–2728, 2015.

[77] H. Yuan, S. C. Kramer, E. H. Lau, B. J. Cowling, and W. Yang. Modeling influenza seasonality in the tropics and subtropics. PLoS Computational Biology, 17(6):e1009050, 2021.

[78] A. Zapranis and A. Alexandridis. Modelling the temperature time-dependent speed of mean reversion in the context of weather derivatives pricing. Applied Mathematical Finance, 15(4):355–386, 2008.

[79] B. Zhou, D. Jiang, B. Han, and T. Hayat. Threshold dynamics and density function of a stochastic epidemic model with media coverage and mean-reverting Ornstein–Uhlenbeck process. Mathematics and Computers in Simulation, 196:15–44, 2022.

[80] G. Zhu, L. Li, Y. Zheng, X. Zhang, and H. Zou. Forecasting influenza based on autoregressive moving average and holt-winters exponential smoothing models. Journal of Advanced Computational Intelligence and Intelligent Informatics, 25(1):138–144, 2021.

